# Community-Level Wastewater Surveillance of Nicotine Biomarkers Tracks Cardiovascular Disease Burden and Myocardial Infarction

**DOI:** 10.64898/2026.10.22.26366217

**Authors:** Katherine M. Joseph, Xuan Qin, Katelyn M. Clark, Feng Li, Emily Hartmann, Juan Nanez, Lorissa Humble, Catherine Turlington, John Balliew, Craig Cassidy, Jagat Narula, Kristina D. Mena, Fuqing Wu

## Abstract

**BACKGROUND:** Tobacco smoking is the largest modifiable contributor to cardiovascular disease (CVD), yet population-level surveillance still relies on self-reported and retail data, which are subject to measuring bias and lag. Urinary and salivary cotinine quantify individual tobacco exposure, but no analogous, objective metric exists at the community level. Wastewater-based epidemiology (WBE) has transformed surveillance of infectious diseases and illicit drug use; whether the same approach can yield a community-level signal of CVD risk is unknown.

**OBJECTIVE:** The primary objective of this study is to track community-level wastewater nicotine biomarkers and evaluate potential correlations with relative CVD burden in El Paso, Texas.

**METHODS:** We integrated wastewater nicotine biomarker surveillance with electronic health record (EHR) data across four sewersheds in El Paso, Texas. Twenty-four-hour composite influent samples were collected monthly (June 2024–May 2025; n=48) and quantified by liquid-chromatography tandem mass spectrometry for five nicotine metabolites. Cotinine, nornicotine, and trans-3’-hydroxycotinine (3HC) were detected in 91.6–100% of samples, whereas tobacco-specific nitrosamines were below limit of detection. CVD cases (January 2021–December 2024) were obtained from the Paso del Norte Health Information Exchange and mapped to sewersheds by residential ZIP code using ArcGIS. At the sewershed level, population-normalized nicotine mass loads tracked the proportional burden of atherosclerosis/ischemia/infarction and heart failure. Across the sewersheds, monthly 3HC mass load and the nicotine metabolite ratio (NMR) were strongly and specifically associated with incident ST-elevation myocardial infarction (STEMI) (3HC vs. STEMI: *ρ* = 0.893, *p* = 0.012), without parallel associations for non-ST-elevation or old myocardial infarction.

**CONCLUSIONS:** To our knowledge, this is the first demonstration that integrating wastewater nicotine surveillance with EHR data can reveal community-level 3HC and NMR as specific indicators of incident STEMI. Our findings indicate wastewater surveillance as a novel, objective tool to inform cessation policy and CVD prevention.

*Lay Words:*

- This study investigated using wastewater to measure population-level nicotine consumption and to see if it was related to local cardiovascular disease.
- The study took place in El Paso, Texas, USA across 4 different wastewater treatment collection points over the course of one year.
- Three metabolites of human nicotine consumption were detected in wastewater samples. The increased nicotine mass loads were proportional with community-level heart failure and atherosclerosis.
- Wastewater can be a helpful tool to inform cardiovascular disease prevention efforts and smoking cessation practices.

## Introduction

Cardiovascular disease (CVD) remains the leading cause of death in the United States.(1,2) It accounted for an estimated 4.69 million hospitalizations and roughly $108 billion in direct medical costs in 2021, with projected costs expected to exceed $130 billion by 2030.(3) Coronary artery disease (CAD), aortic atherosclerosis, cerebrovascular disease, and peripheral artery disease share a common pathophysiologic substrate of endothelial injury, oxidative stress, and accelerated atherogenesis, and converge clinically on myocardial infarction (MI), heart failure, stroke, and arrhythmia.(4–7) MI is further classified based on electrocardiographic findings into ST-elevation MI (STEMI), which results from complete coronary artery occlusion and non-ST-elevation MI (NSTEMI), which typically arises from a partial occlusion.(8) Among modifiable risk factors, such as hypertension, dyslipidemia, obesity, diabetes, and tobacco use, cigarette smoking remains the single largest preventable contributor to cardiovascular morbidity and mortality.(7,9)

Smoking raises the risk of CAD by approximately 30% and is causally implicated in MI, arrhythmias, heart failure, stroke, and sudden cardiac death.(9,10) Mechanistically, tobacco-derived reactive oxygen species drive endothelial dysfunction, vascular inflammation, lipid oxidation, and platelet activation, providing a coherent biological pathway from exposure to acute coronary events.(9,11–13) Cessation reduces CVD risk within five years, even amongst heavy smokers, which makes accurate population-level monitoring of tobacco exposure a high-yield target for cardiovascular prevention.(10,13) However, the contemporary tobacco landscape is increasingly heterogeneous; combustible cigarette use has declined while heated-tobacco, smokeless, and electronic-cigarette use has expanded, with mounting evidence of independent cardiovascular harm.(14–22) Self-reported metrics and retail sales data are poorly suited to capture these rapid shifts, motivating objective measures of community nicotine exposure with a decreased ascertainment bias.

Absorbed nicotine is oxidized in the liver, predominantly by CYP2A6, with 70–80% converted to cotinine and approximately 40–50% of cotinine subsequently hydroxylated to trans-3’-hydroxycotinine (3HC) (**Figure 1A**).(23,24) Minor pathways yield nornicotine via N-demethylation and the tobacco-specific nitrosamines N’-nitrosonornicotine (NNN) and 4-(methylnitrosamino)-1-(3-pyridyl)-1-butanone (NNK), which are both classified as Group 1 human carcinogens.(25–29) At the individual level, urinary or salivary cotinine, 3HC, and the 3HC/cotinine nicotine metabolite ratio (NMR)-a validated phenotypic proxy for CYP2A6 activity and the intensity of nicotine intake - are well-established biomarkers of tobacco exposure and metabolic phenotype.(30) Several have been recovered intact from municipal wastewater sources.(31,32)

**Figure 1:**
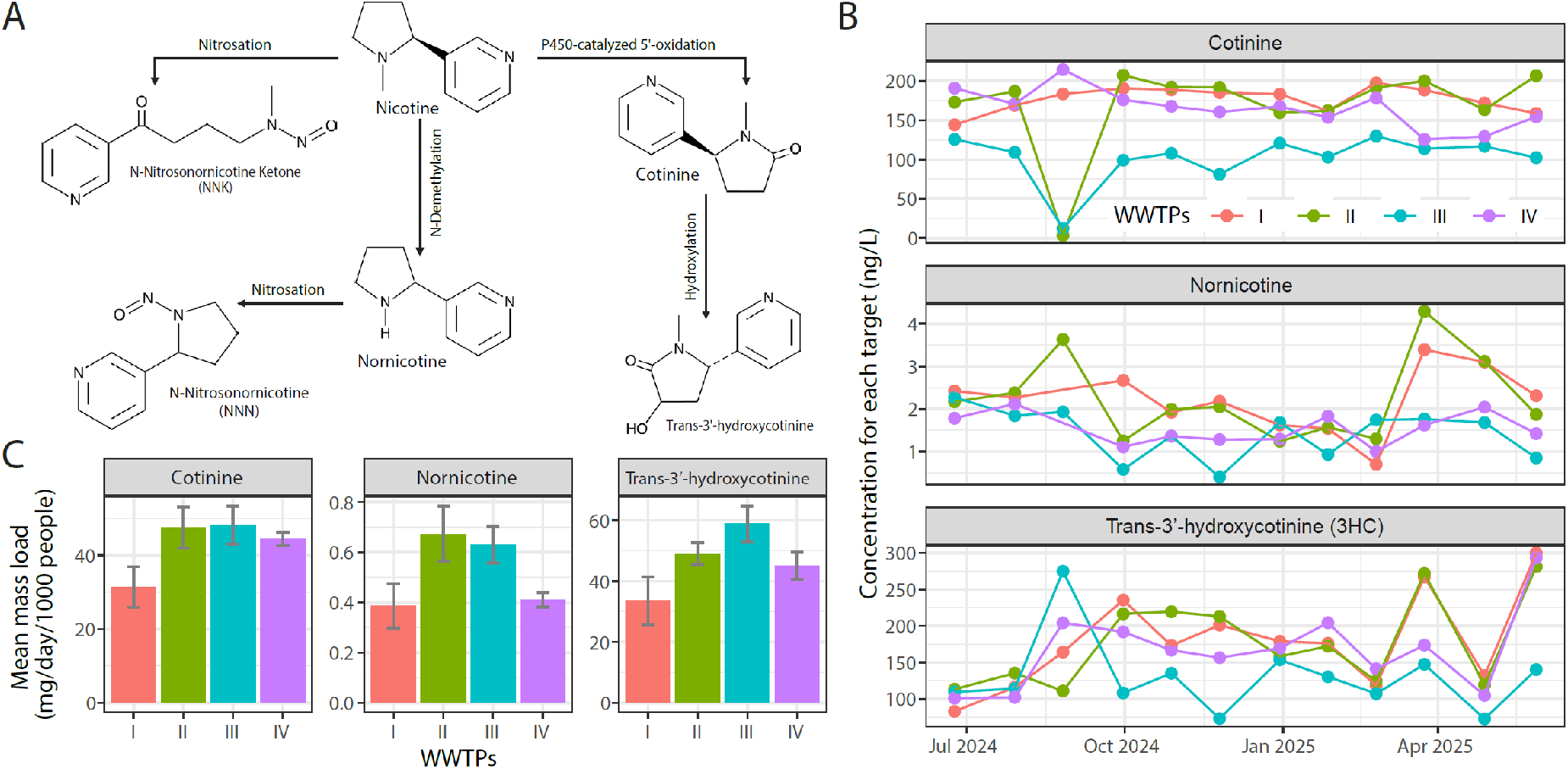
Wastewater-based surveillance of nicotine biomarkers across four sewersheds in the City of El Paso. (**A**) Pharmacokinetic pathways of nicotine metabolism in the human body. Nicotine is primarily converted to cotinine via P450-catalyzed oxidation, which is further hydroxylated to trans-3’-hydroxycotinine (3HC). Nitrosation of nornicotine and nicotine produces the tobacco-specific nitrosamines N-nitrosonornicotine (NNN) and N-nitrosonornicotine ketone (NNK), respectively. (**B**) Temporal trends in wastewater concentrations (ng/L) of cotinine, nornicotine, and 3HC measured monthly across four sewersheds (WWTP I-IV) from June 24, 2024 - May 28, 2025. (**C**) Mean mass load of Cotinine, Nornicotine, and 3HC across the four sewersheds over the study period. Error bars represent standard error, and WWTPs are color-coded consistently across all panels (I: red, II: green, III: teal, IV: purple).

Wastewater-based epidemiology (WBE) leverages the small-molecule and pathogen biomarkers excreted by entire populations converging at wastewater treatment plants (WWTPs), generating a non-invasive, near real-time, geographically resolved measure of community exposure free of recall, participation, and stigma biases.(33–35) Two precedents anchor the concept. First, ambient PM2.5 and ozone monitoring transformed cardiovascular epidemiology of air pollution by providing objective, spatially resolved exposure estimates that closely track day-to-day variation in MI and stroke admissions.(36–38) Second, wastewater quantification of SARS-CoV-2, poliovirus, and mpox virus now anchors infectious disease surveillance worldwide by tracking circulating viral loads as a marker of community endemicity and impending clinical waves.(39–42) Wastewater chemistry can deliver an analogous community-scale exposure signal for tobacco. Prior WBE studies have used cotinine, 3HC, and anabasine to quantify temporal and spatial trends in nicotine consumption, distinguish combustible from non-combustible products, and triangulate against tobacco sales records.(31,43–50) These efforts have remained mostly descriptive. To our knowledge, no prior study has linked wastewater-derived nicotine biomarkers to clinical cardiovascular outcomes in the same geographic catchment. This omission is consequential, because individual-level cotinine and NMR are established correlates of incident coronary events, yet population-health systems still lack an objective, scalable analog for monitoring tobacco-attributable cardiovascular risk at the community level in near real time.(51–56)

The primary objective of this study was to track community-level wastewater nicotine biomarkers and evaluate potential correlations with relative burden of CVD in El Paso, Texas. The city is served by four WWTPs covering approximately 87% of El Paso County. We integrated 12 months of LC-MS/MS quantification of five nicotine metabolites in monthly composite wastewater samples (June 2024 - May 2025) with electronic-health-record (EHR)– derived cardiovascular diagnoses to address three questions.(57,58) First, whether nicotine metabolites are reliably detectable in municipal wastewater across a city. Second, whether population-normalized metabolite mass loads correlate with the relative burden of specific cardiovascular conditions across sewersheds. Third, whether monthly fluctuations in wastewater nicotine track incident acute coronary events, in particular STEMI.

## Materials and Methods

### Wastewater sample collection

Wastewater samples were collected, processed, and quantified using protocols described previously in our study detecting other biomarkers of drugs in wastewater.(59) Twenty-four-hour, time-weighted composite samples of raw influent were collected from four WWTPs in El Paso, Texas (deidentified throughout as WWTPs I–IV) from June 24, 2024 to May 28, 2025. The four plants are operated by El Paso Water Utilities and collectively serve approximately 751,982 residents (≈87% of El Paso County); plant-specific service populations were approximately 98,505 for WWTP I, 125,235 for WWTP II, 127, 235 for WWTP III, and 399,424 for WWTP IV.(39,40) Wastewater sample daily flow rates were reported by each wastewater facility (**Table S1**). HACH AS950 all-weather refrigerated samplers collected wastewater aliquots every 30 minutes over each 24-hour period. Composite samples (500 mL) were shipped overnight on ice to the laboratory in Houston and stored at –20 °C until testing. All samples were collected on Mondays, by agreement with the participating utilities, to minimize day-of-week variability in across-plant comparisons

### Sample Preparation and Quantification with LC-MS/MS

Sample processing followed our previously described method.(59) Briefly, raw wastewater (40 mL) was centrifuged and then further filtered via 0.22 µm polyethersulfone membrane (Millipore Sigma, #SCGP00525). The filtrate (30 mL) was spiked with deuterated internal standards (100 ng/L each of cotinine-d3; Cayman Chemical) prior to filtration. Solid-phase extraction (SPE) was performed on Oasis MCX cartridges pre-conditioned sequentially with HPLC-grade methanol, HPLC-grade water, and 0.1% (v/v) HCl in HPLC-grade water. After sample loading and bottle rinses with HPLC-grade water and 0.1% HCl were added back onto each cartridge, cartridges were washed with methanol, dried under vacuum, and then air-dried at room temperature.

Analytes were eluted with 4 mL of tert-butyl methyl ether (MTBE) /isopropanol/ammonium hydroxide (78:20:2, v/v/v; Fisher Scientific, #E127). Eluates were collected in 5 mL tubes, evaporated under a gentle nitrogen stream at room temperature, reconstituted in 150 µL of 50% aqueous methanol, vortexed, and centrifuged at 15,000 rcf for 15 minutes. The SPE workflow concentrated each sample 200-fold (30 mL → 150 µL).

Five microliters of supernatant were injected into a Thermo Vanquish UHPLC system coupled to a Thermo TSQ Quantis tandem mass spectrometer (Thermo Fisher Scientific). Analytes and internal standards (IS) were separated on an Acquity BEH-C18 column (2.1 × 100 mm, 1.7 µm; Waters, Milford, MA) using a 5%–95% organic gradient over a 20-minute run with mobile phases of water (10 mM ammonium formate) and acetonitrile (0.1% formic acid, v/v) at 0.3 mL/min and 40 °C. Retention times and selected reaction monitoring (SRM) ion transitions for each analyte and IS are reported in **Table S2**. Cotinine-d3 served as the deuterated internal standard because its chromatographic and ionization behaviors closely matched those of the five target nicotine metabolites in the wastewater matrix, minimizing ion-suppression effects and improving quantitative accuracy; recoveries are reported in **Table 1**. Analytes were monitored in SRM mode under positive electrospray ionization with a 3,500 V spray voltage. Ultra-high-purity nitrogen served as sheath, auxiliary, and sweep gases (50, 10, and 1 arbitrary unit, respectively); ultra-high-purity argon served as collision gas. Ion-transfer tube and vaporizer temperatures were 325°C and 350°C, respectively, and signals were recorded for 0.5-17 minutes per injection.

**Table 1:** Method validation parameters and extraction recovery rates.

| Targets | LOD<br>(ng/L) | LLOQ<br>(ng/L) | Linear<br>range<br>(ng/L) | Nominal<br>concentration (ng/L) | Peak area |  | Recovery<br>(%) |
| --- | --- | --- | --- | --- | --- | --- | --- |
|  |  |  |  |  | SPE-ed | Post SPE |  |
| Cotinine | 1 | 3 | 3 – 800 | 10 | 50060 | 138171 | 36.23 |
|  |  |  |  | 100 | 557926 | 1358720 | 41.06 |
|  |  |  |  | 700 | 4564610 | 9941255 | 45.92 |
| Trans-3'-<br>hydroxycot<br>inine | 1 | 3 | 3-800 | 10 | 13972 | 22812 | 61.25 |
|  |  |  |  | 100 | 148340 | 212160 | 69.92 |
|  |  |  |  | 700 | 929205 | 2088146 | 44.50 |
| Nornicotin<br>e | 0.1 | 0.25 | 0.25-10 | 0.5 | 4276 | 8181 | 52.26 |
|  |  |  |  | 2 | 16108 | 31288 | 51.48 |
|  |  |  |  | 8 | 62142 | 148902 | 41.73 |
| NNN | 0.25 | 0.5 | 0.5-10 | 1 | 7013 | 16368 | 42.84 |
|  |  |  |  | 2 | 13649 | 31743 | 43.00 |
|  |  |  |  | 8 | 64121 | 147885 | 43.36 |
| NNK | 3 | 10 | 10-800 | 30 | 8055 | 13444 | 59.92 |
|  |  |  |  | 100 | 25608 | 50230 | 50.98 |
|  |  |  |  | 700 | 158925 | 285388 | 55.69 |
| Cotinine-d3 | \ | \ | \ | 100 | 842300 | 1566063 | 53.78 |

Six-level calibration curves were generated by spiking HPLC-grade water with certified reference standards over the analyte ranges observed in wastewater: 3–800 ng/L for cotinine and 3HC, 0.25–10 ng/L for nornicotine, 0.5–10 ng/L for NNN, and 10–800 ng/L for NNK (concentrations refer to standards spiked into water prior to SPE). HPLC-grade water blanks served as negative controls. Method validation followed U.S. Food and Drug Administration (FDA) bioanalytical guidance and included assessment of linearity, limits of detection (LOD; signal-to-noise ratio of 3:1) and lower limits of quantitation (LLOQ; signal-to-noise ratio of 10:1), SPE recovery, and intra-and inter-day precision and accuracy were measured at three quality control (QC) levels (**Tables 1–3**).(60) SPE recovery was calculated as 100% x (peak area of analyte spiked into water before SPE) / (peak area of analyte spiked into post-SPE eluted HPLC-grade water at the matched nominal concentration); the same calculation was applied to the deuterated internal standards. Intra-day precision and accuracy were determined from triplicate measurements of three QC levels prepared and analyzed on the same day (**Table 2**); inter-day precision and accuracy were determined from triplicate measurements of the same three QC levels analyzed across three independent runs on different days (**Table 3**).

**Table 2.** Intra-day Precision and Accuracy.

|  | **Nominal**<br>**concentration**<br>**(ng/L)** | **Test 1**<br>**(ng/L)** | **Test 2**<br>**(ng/L)** | **Test 3**<br>**(ng/L)** | **Average**<br>**(ng/L)** | **Precision**<br>**(%)** | **Accuracy (%)** |
| --- | --- | --- | --- | --- | --- | --- | --- |
| Cotinine | 10 | 10.04 | 8.98 | 8.49 | 9.17 | 8.67 | -8.29 |
|  | 100 | 106.28 | 98.41 | 92.22 | 98.97 | 7.12 | -1.03 |
|  | 700 | 716.04 | 717.83 | 702.32 | 712.07 | 1.19 | 1.72 |
| Trans-3'-<br>hydroxycotin<br>ine | 10 | 11.23 | 10.25 | 8.56 | 10.01 | 13.49 | 0.14 |
|  | 100 | 108.36 | 123.22 | 93.84 | 108.47 | 13.54 | 8.47 |
|  | 700 | 726.38 | 707.13 | 643.93 | 692.48 | 6.23 | -1.07 |
| Nornicotine | 0.5 | 0.56 | 0.54 | 0.58 | 0.56 | 3.84 | 11.73 |
|  | 2 | 1.90 | 2.07 | 2.03 | 2.00 | 4.37 | 0.04 |
|  | 8 | 6.97 | 8.43 | 8.30 | 7.90 | 10.27 | -1.24 |
| NNN | 1 | 1.02 | 0.99 | 1.02 | 1.01 | 1.38 | 0.93 |
|  | 2 | 1.99 | 1.89 | 2.06 | 1.98 | 4.16 | -1.01 |
|  | 8 | 7.51 | 8.59 | 8.72 | 8.27 | 8.04 | 3.43 |
| NNK | 30 | 30.17 | 31.37 | 30.79 | 30.78 | 1.95 | 2.60 |
|  | 100 | 112.46 | 92.97 | 100.89 | 102.10 | 9.60 | 2.10 |
|  | 700 | 679.34 | 704.10 | 702.42 | 695.29 | 1.99 | -0.67 |

**Table 3.** Inter-day Precision and Accuracy.

|  | Nominal<br>concentration<br>(ng/L) | Test 1<br>(ng/L) | Test 2<br>(ng/L) | Test 3<br>(ng/L) | Average<br>(ng/L) | Precision<br>(%) | Accuracy<br>(%) |
| --- | --- | --- | --- | --- | --- | --- | --- |
| Cotinine | 10 | 9.44 | 9.55 | 9.17 | 9.39 | 2.12 | -6.13 |
|  | 100 | 103.79 | 103.48 | 94.34 | 100.54 | 5.34 | 0.54 |
|  | 700 | 722.12 | 689.10 | 651.66 | 687.63 | 5.13 | -1.77 |
| Trans-3'-<br>hydroxycotin<br>ine | 10 | 8.52 | 9.48 | 10.17 | 9.39 | 8.83 | -6.11 |
|  | 100 | 106.90 | 104.76 | 105.83 | 105.83 | 1.01 | 5.83 |
|  | 700 | 733.52 | 704.24 | 754.24 | 730.66 | 3.44 | 4.38 |
| Nornicotine | 0.5 | 0.57 | 0.57 | 0.51 | 0.55 | 6.71 | 10.16 |
|  | 2 | 1.93 | 2.02 | 1.78 | 1.91 | 6.45 | -4.40 |
|  | 8 | 7.93 | 8.24 | 7.86 | 8.01 | 2.52 | 0.17 |
| NNN | 1 | 0.89 | 1.01 | 0.92 | 0.94 | 6.48 | -5.81 |
|  | 2 | 1.91 | 2.14 | 1.82 | 1.96 | 8.47 | -2.06 |
|  | 8 | 8.51 | 8.23 | 8.37 | 8.37 | 1.63 | 4.62 |
| NNK | 30 | 33.36 | 26.77 | 28.57 | 29.57 | 11.52 | -1.44 |
|  | 100 | 104.59 | 112.76 | 96.17 | 104.51 | 7.94 | 4.51 |
|  | 700 | 680.18 | 655.63 | 676.42 | 670.74 | 1.97 | -4.18 |

### Calculation of drug mass loads and per-capita consumption

Daily mass loads of cotinine, nornicotine, and 3HC were calculated by multiplying each measured concentration (ng/L) by the corresponding daily influent flow volume for the sampled WWTP (**Equation 1**).(61–63) Daily influent flow data (million gallons per day, MGD) were provided by each WWTP. Mass loads were then normalized to sewershed population to yield per-capita estimates expressed as mg of analyte per day per 1,000 served residents (**Equation 2**). Per-capita estimates were not back-calculated to nicotine intake; they do not account for analyte-specific urinary and fecal excretion fractions, which differ markedly across cotinine, 3HC, and nornicotine. We report mass loads rather than reconstructed consumption to avoid compounding excretion-rate and in-sewer-stability uncertainties.(64,65) All numerical data has been rounded to the nearest integer as dictated by the propagation of uncertainty.

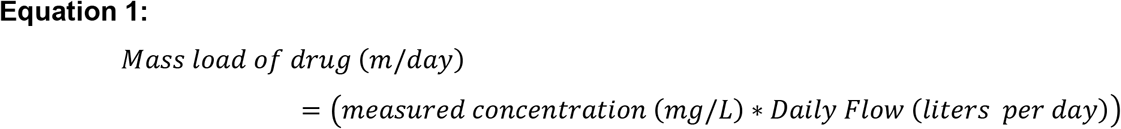

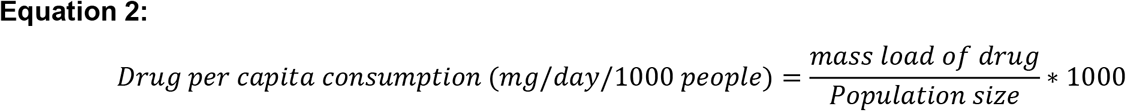

### Patient data abstraction related to cardiovascular disease

Patient-level cardiovascular case records were obtained from the Paso del Norte Health Information Exchange (PHIX), a regional nonprofit health information exchange that aggregates and deidentifies clinical data from 11 major hospitals serving El Paso, Texas. Records spanning January 2021 through December 2024 (36 months) were extracted for patients with a healthcare encounter related to at least 1 ICD-10 diagnosis code of interest (**Table S3**) and excluding any patients with an end stage renal disease diagnosis (ICD-10 code N18.6*) or a cancer diagnosis (ICD-10 codes C00-C96) to avoid confounding health conditions. Only the first recorded diagnosis per patient within each diagnostic category was retained, to capture incident rather than recurrent cases. Consistent with the PHIX disclosure-control policy, any diagnosis-by-ZIP code (or diagnosis-by-ZIP code-by-month) cell containing fewer than 10 patients was suppressed at the source and not released for analysis; such suppressed cells are noted where they constrain interpretation.

Each qualifying patient encounter was assigned a unique de-identified study identifier prior to release. Demographic variables included age, sex, race and ethnicity, marital status, and residential ZIP code. Patients were assigned to one of the four sewersheds based on residential ZIP code: ZIP codes falling entirely within a single sewershed were mapped directly, and the two ZIP codes whose boundaries crossed sewershed lines were assigned to the sewershed covering ≥75% of their geographic area. Spatial mapping was performed in ArcGIS Pro version 3.6.3 (Esri, Redlands, CA) using utility-provided sewershed shapefiles and U.S. Census Bureau ZIP Code Tabulation Area shapefiles. CVD outcomes were classified by ICD-10 diagnostic codes covering abnormal heart rhythm, atherosclerosis/ischemia/infarction, chest pain, heart failure, hypertension, nicotine dependence, valve disorders, atrial fibrillation/flutter, and syncope/collapse; specific code ranges are listed in **Table S3**. The proportion of patients in each ICD-10 diagnostic category was calculated by dividing the number of patients with the respective diagnosis over the total population from the dataset who resided in that sewershed.

### 2.5. Statistical analysis

Wastewater nicotine biomarker mass loads were compared across the four WWTP locations for each metabolite using Kruskal-Wallis test. Spearman rank correlation coefficients (ρ) were used to evaluate associations between wastewater nicotine biomarker mass loads and cardiovascular outcomes. Three pre-specified analyses were performed: (i) cross-sewershed associations between the mean wastewater mass load of each biomarker and the proportion of CVD patients in each diagnostic category across the four sewersheds; (ii) city-level monthly associations between city-aggregated biomarker mass load and counts of incident STEMI subtypes during the overlapping wastewater-EHR period (June–December 2024, n = 7 months); and (iii) a secondary analysis using mean monthly incident STEMI counts averaged across the full 2021– 2024 EHR window (n = 12 calendar months). Two-sided p-values were considered significant at α = 0.05; both ρ and p-values are reported throughout. Additionally, we indicate where associations would survive a Bonferroni correction for the four biomarker tests within each diagnostic category. All analyses were performed in R version 4.4.1 using cor.test() with method = “spearman”; figures were generated with ggplot2 and ComplexHeatmap.

The sample size was determined by sample availability rather than a priori power calculation. A sensitivity power analysis indicated that with an n = 48, this study had a 80% power to detect correlations of |r| ≥ 0.39 (α = 0.05) in a simple linear regression. This effect size threshold was applied uniformly to all analyses performed. Observed effect sizes were generally at or above this threshold, indicating that the study was adequately powered to detect associations identified as statistically significant.

## Results

### Analytical method validation

The LC-MS/MS method was validated against U.S. FDA bioanalytical guidance for all five target analytes (cotinine, nornicotine, 3HC, NNN, and NNK).(60) HPLC-grade water blanks showed no detectable peaks at the retention times of the targets, confirming method specificity. LODs were 1 ng/L for cotinine and 3HC, 0.1 ng/L for nornicotine, 0.25 ng/L for NNN, and 3 ng/L for NNK, with corresponding LLOQs of 3, 0.25, 0.5, and 10 ng/L (**Table 1**). Linear calibration ranges spanning the concentrations observed in El Paso wastewater were established for each analyte, with R^2^ ≥ 0.99 throughout (**Figure S1**). SPE recovery across the three QC levels ranged from 36.2–45.9% for cotinine, 41.7–52.3% for nornicotine, 44.5–69.9% for 3HC, 42.8–43.4% for NNN, and 51.0–55.7% for NNK; recovery for the cotinine-d3 internal standard was 53.8% (**Table 1**). Intra-day precision and accuracy ranged from 1.19–13.54% and-8.29–11.73% across all analytes (**Table 2**), and inter-day precision and accuracy from 1.01–11.52% and 6.13–10.16% (**Table 3**). All performance metrics remained within the ±15% acceptance window prescribed by FDA guidance, supporting reliable quantitative performance for wastewater nicotine biomarker monitoring.(60)

### Profile of community nicotine exposure across the city

Applying the validated method, we analyzed monthly composite wastewater from each of the four El Paso WWTPs over a 12-month surveillance window (June 2024–May 2025; n = 48). Cotinine, nornicotine, and 3HC were detectable in 91.6–100% of samples across all four sewersheds (**Figure S1**), whereas the tobacco-specific nitrosamines NNN and NNK were below LOD in every sample, consistent with their very low fractional excretion relative to the major nicotine metabolites.(30,66,67) Subsequent analyses therefore focus on cotinine, nornicotine, and 3HC as community-level indicators of nicotine exposure.

Wastewater concentrations of all three biomarkers were generally stable over the 12-month window, with month-to-month variation smaller than between-plant differences (**Figure 1B**). Cotinine and 3HC concentrations were comparable in magnitude (153.6 ± 45.4 ng/L and 163.6 ± 59.8 ng/L, respectively) and tracked one another temporally across plants; nornicotine concentrations were approximately two orders of magnitude lower (1.8 ± 0.8 ng/L), consistent with its minor role in nicotine metabolism. Population-normalized mass loads, calculated using plant-specific influent flow and service-area population, revealed clear spatial heterogeneity in community nicotine exposure (**Figure 1C**). WWTP-I had the lowest mass loads across all three analytes (cotinine 31 ± 6 mg/day/1,000 people; 3HC 34 ± 8 mg/day/1,000 people; nornicotine 400 ± 90 µg/day/1,000 people), while WWTPs II and III had the highest (cotinine up to 48 ± 5; 3HC up to 59 ± 6; nornicotine up to 700 ± 100), an approximately 1.5-to 1.8-fold gradient in community nicotine exposure within the city. Overall, group differences across the WWTPs were significant for nornicotine (*Kruskal-Wallis p* = 0.043) and 3HC (*Kruskal-Wallis p* = 0.040), but not for cotinine (*p*=0.141).

### Higher community nicotine exposure is associated with higher CVD disease burden

To examine whether this spatial gradient in nicotine exposure maps onto CVD disease burden, we extracted incident CVD diagnoses from PHIX and assigned each patient to one of the four sewersheds based on their residential ZIP code. All patients who lived outside of sewersheds covered by WWTPs I – IV, were excluded. Patient demographics by sewershed are summarized in **Figure S2**: 76.7% of patients were ≥40 years of age (the conventional age threshold for elevated CVD risk), 74.0% identified as Hispanic, 85.9% as White, 54.0% as female, and marital status was approximately evenly distributed across never married (27.5%), married (27.2%), and unknown (33.3%) categories. We retained the full age range in the primary analyses, since young adults can still contribute to nicotine consumption.(68)

Because EHR data captured diagnosed and treated cases rather than the underlying community disease burden, we computed the proportion of patients in each ICD-10 diagnostic category relative to the total CVD patient population within each sewershed. This condition-specific proportion compares the relative composition of CVD across catchments with differing nicotine exposure, while remaining robust to absolute differences in catchment size or care-seeking behaviors. Spearman rank correlations were then computed between mean nicotine biomarker mass loads and condition-specific proportions across the four sewersheds.

Sewersheds with higher nicotine mass loads consistently exhibited higher proportions of CVD patients across multiple diagnostic categories (**Figure 2A**). The strongest and most consistent positive associations were observed for heart failure and atherosclerosis/ischemia/infarction (ρ = 1.0 for both cotinine and 3HC; ρ = 0.8 for nornicotine). Tobacco/nicotine-related diagnoses were also strongly correlated, particularly with nornicotine (ρ = 1.0). Moderate positive associations were observed for valve disorders, and atrial fibrillation/flutter (ρ = 0.6–0.8), with weaker correlations for hypertension and abnormal heart rhythm (ρ = 0.2–0.4). Chest pain showed a negative correlation with cotinine and 3HC (ρ = –0.8), which likely reflects diagnostic substitution. In sewersheds with a higher burden of confirmed coronary disease, undifferentiated chest pain accounts for a smaller share of the CVD case mix. Scatter plots illustrate the underlying relationships for heart failure (**Figure 2B**) and atherosclerosis/ischemia/infarction (**Figure 2C**), with WWTP-I (lowest nicotine loads) anchoring the low end and WWTP-III the high end of both gradients. Together, these results demonstrate a consistent positive association between community-level nicotine exposure and the relative burden of specific cardiovascular conditions across sewersheds within the city of El Paso.

**Figure 2:**
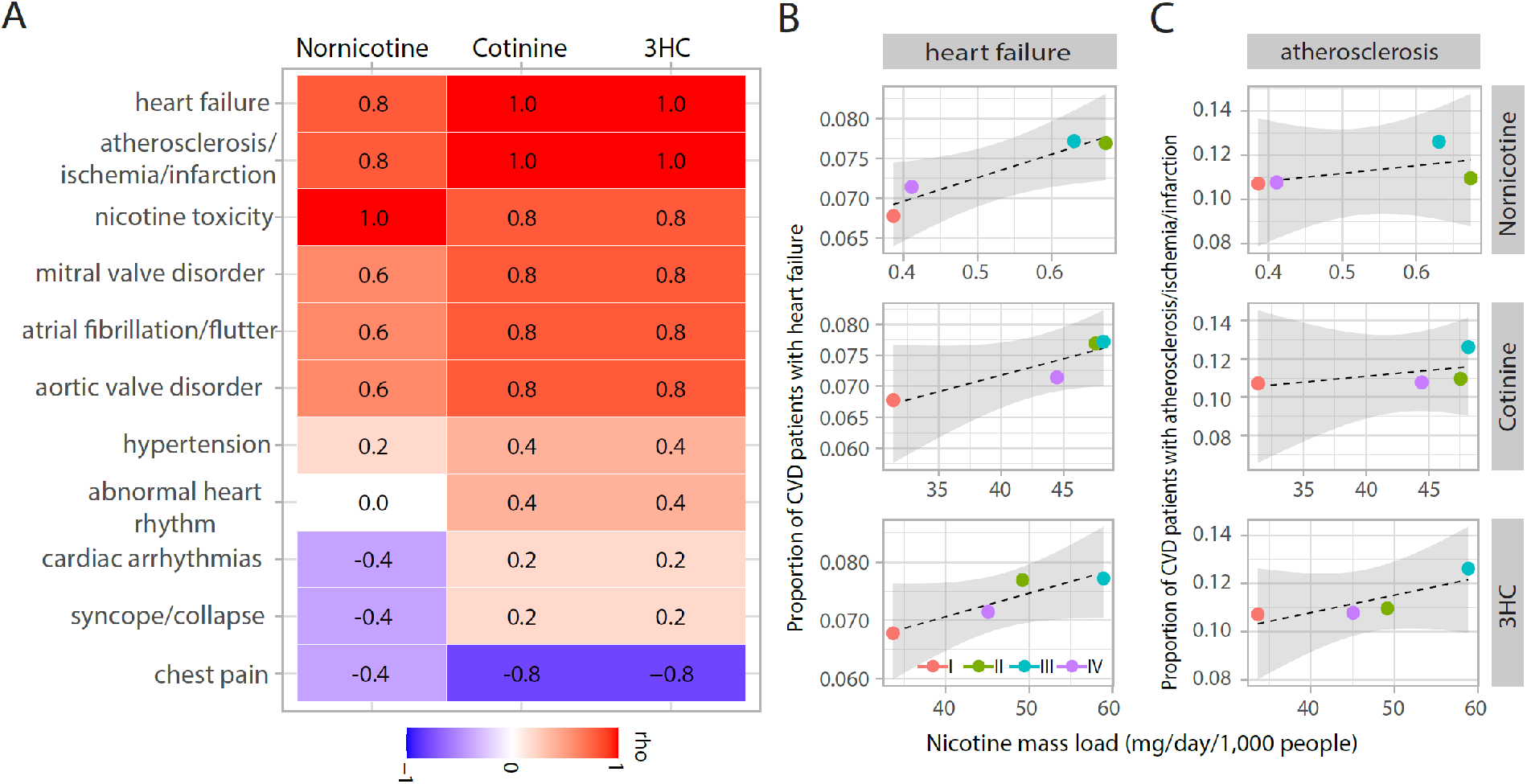
Association between wastewater nicotine biomarkers and CVD disease burden across the four sewersheds. (**A**) Heatmap of Spearman correlation coefficients (ρ) between mean nicotine biomarker mass loads (nornicotine, cotinine, and 3HC) and the mean monthly proportion of CVD patients across eleven diagnosis categories. Wastewater biomarker data (June 2024 - May 2025) and CVD diagnosis records (January 2021-December 2024) were each aggregated by WWTP catchment area (n = 4). Color bar indicates the coefficient ρ, with red indicating positive and blue indicating negative associations. (**B-C**) Scatter plots showing the association between mean nicotine mass load (mg/day/1,000 people) and mean monthly proportion of CVD patients for (**B**) heart failure and (**C**) atherosclerosis/ischemia/infarction, faceted by nicotine biomarker (nornicotine, cotinine, and 3HC). Each point represents one WWTP site (I-IV, color-coded), with dashed regression lines and shaded 95% confidence intervals.

### Monthly 3HC mass load and nicotine metabolite ratio are significantly associated with STEMI at the city level

The sewershed-level analysis identified atherosclerosis/ischemia/infarction as the diagnostic category most strongly associated with community nicotine exposure. To resolve this association at a finer diagnostic and temporal scale, we examined whether monthly fluctuations in city-level nicotine exposure track variation in incident STEMI, the most acute and pathobiological proximate manifestation within that category. Monthly nicotine mass loads were averaged across the four WWTPs to derive city-level monthly exposure estimates and correlated with monthly new STEMI diagnosis counts over the overlapping wastewater–EHR window (June–December 2024, n = 7 months). In addition to the three individual biomarkers, we evaluated the nicotine metabolite ratio (NMR; 3HC/cotinine) as a population-level proxy for nicotine metabolism. A higher NMR indicates faster CYP2A6-mediated nicotine clearance, which at the individual level has been associated with more intensive smoking behavior, higher daily cigarette consumption, and reduced success of nicotine-replacement cessation therapy.(51–53)

Among the 4-exposure metrics examined, 3HC and NMR showed the strongest and most consistent positive associations with STEMI incidence (**Figure 3A**). For inferior-wall STEMI (I21.19), 3HC was strongly associated (ρ = 0.93, *p* = 0.003), followed by NMR (ρ = 0.85, *p* < 0.05); for unspecified STEMI (I21.3), 3HC again showed a significant positive association (ρ = 0.82, P < 0.05). Combining STEMI subtypes, monthly 3HC mass load remained strongly correlated with total incident STEMI (ρ = 0.893, *p* = 0.012; **Figure 3B**). Temporal-trend analysis revealed visual concordance between monthly 3HC mass load and total MI diagnosis counts across the surveillance window, with co-occurring peaks in August and October 2024 (**Figure 3C**). We performed this comprehensive analysis as a consistent temporal trend was found across the MI subclassifications. Importantly, the biomarker (not NMR) was not strongly associated with non-ST-elevation MI (NSTEMI) or with chronic MI. This pattern of diagnostic specificity is biologically coherent. STEMI most directly reflects acute plaque rupture and coronary thrombosis triggered by active tobacco exposure, whereas NSTEMI and chronic MI have more heterogeneous etiologies that include demand ischemia, microvascular disease, and prior remodeling.(69,70) The selective association with STEMI strengthens confidence that the wastewater signal reflects acute biological mechanisms linking nicotine exposure to coronary events rather than nonspecific confounding.

**Figure 3.**
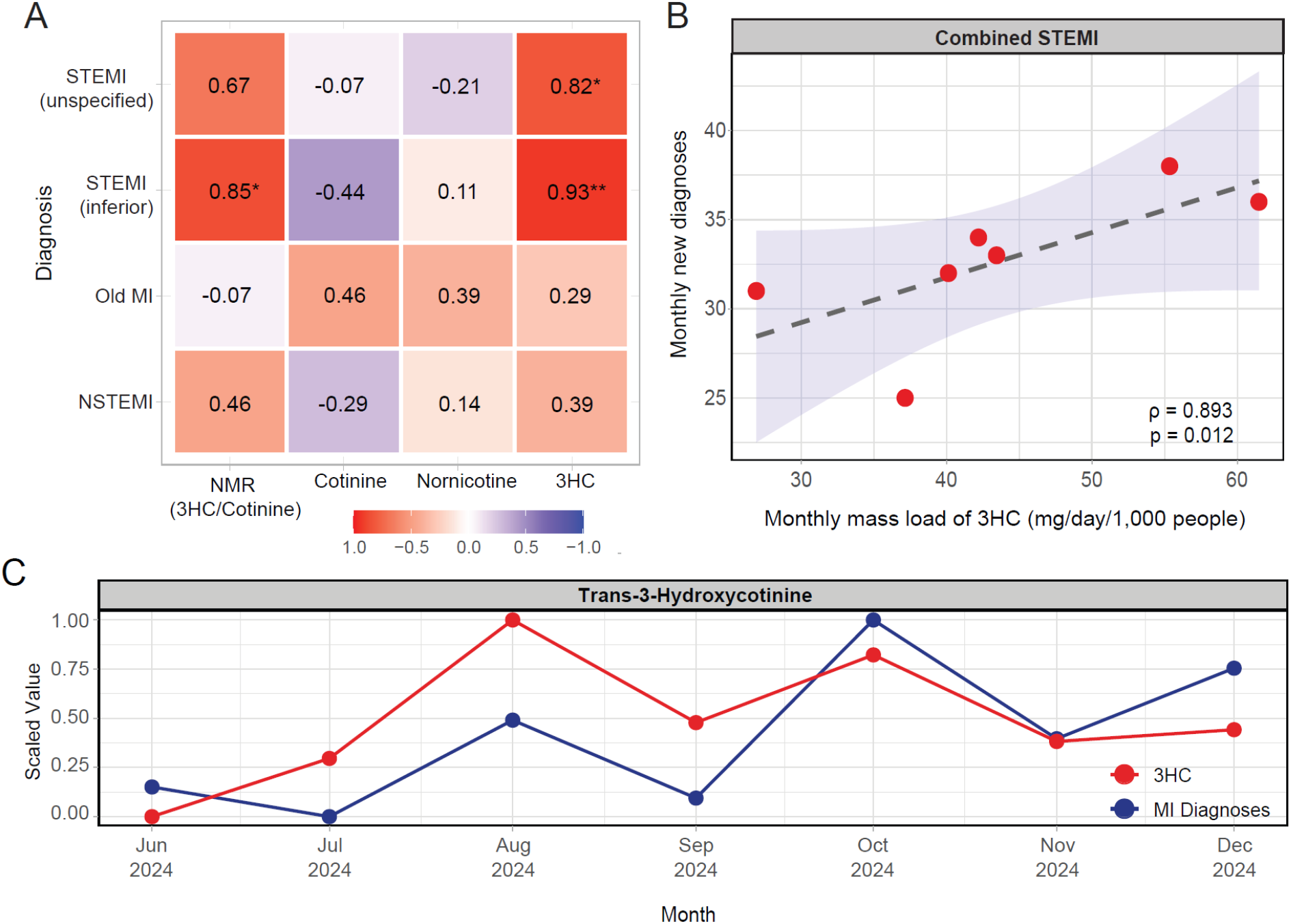
Association between monthly wastewater nicotine biomarker mass loads and MI diagnoses at the city level. **(A)** Heatmap of Spearman correlation coefficients (ρ) between monthly mass loads of four nicotine biomarkers: the nicotine metabolite ratio (NMR, 3HC/cotinine), cotinine, nornicotine, and 3HC, and monthly new diagnoses for four CVD conditions: STEMI inferior (I21.19), STEMI unspecified (I21.3), Old MI (I25.2), and NSTEMI (I21.4). Wastewater biomarker mass loads were averaged across four WWTPs to derive city-level monthly estimates. Significance annotations reflect Spearman correlation adjusted p-values (* p < 0.05, ** p < 0.01). **(B)** Scatter plots showing the association between monthly 3HC mass load (mg/day/1,000 people) and monthly new diagnoses for STEMI (STEMI inferior and STEMI unspecified). Each point represents one calendar month. Dashed lines represent fitted regression lines with shaded 95% confidence intervals. Spearman correlation coefficients (ρ) and p-values are shown in each panel. (**C**) Temporal trends of 3HC mass loads with all myocardial infarction cases (NSTEMI, STEMI, old myocardial infarction) to show correlations from June 2024 to December 2024 (the overlap period). Both data are scaled to 0∼1 on a min max scale for visualization.

A pre-specified secondary analysis using mean monthly STEMI counts averaged across the full 2021–2024 EHR window (n = 12 calendar months) yielded a consistent pattern: 3HC and NMR again showed the strongest associations with inferior STEMI (3HC: ρ = 0.81, *p* < 0.01; NMR: ρ = 0.79, *p* < 0.01), while cotinine and nornicotine, as well as NSTEMI and old MI, remained non-significant (**Figure S3**). Together, these results indicate a robust and biologically specific positive association between community wastewater 3HC and NMR and acute coronary events, suggesting that wastewater-derived nicotine biomarkers can function as community-level indicators of acute CVD burden.

## 4. Discussion

In this study, we demonstrate that community-level wastewater nicotine biomarkers track both the cross-sectional burden and a correlating incidence of CVD. At the sewershed level, population-normalized cotinine, 3HC, and nornicotine mass loads correlated with the proportional burden of atherosclerosis/ischemia/ infarction and heart failure across catchments. Overall, monthly 3HC mass load and the nicotine metabolite ratio were strongly associated with incident STEMI, the most acute and pathobiological proximate cardiovascular endpoint, without parallel associations with NSTEMI or chronic MI. The study did not find much correlation of nornicotine with CVD and did not detect the presence of NNN and NNK in wastewater. To our knowledge, this is the first demonstration that an aggregate, objective wastewater signal of nicotine exposure aligns with both the relative distribution and the timing of acute coronary events in the same geographic catchment.

The findings extend two established paradigms in cardiovascular and population surveillance. The first is the use of wastewater to quantify community-level pathogen circulation. Wastewater quantification of SARS-CoV-2, poliovirus, mpox virus, and other agents is now integral to infectious disease surveillance worldwide because viral loads in influent integrate over an entire catchment, capturing asymptomatic and undiagnosed shedders and leading clinical reporting by days to weeks.(33–35,39–42) Our results suggest that an analogous logic applies to nicotine.

Aggregate excretion of cotinine, 3HC, and nornicotine into the sewer integrates the smoking and non-combustible tobacco behavior of an entire community, decreasing risk of recall, social-desirability, and participation biases that limit self-report metrics, and producing a complementary continuous quantitative signal that can be interpreted as a marker of tobacco-exposure endemicity to assist other preventative public health measures. The second paradigm is environmental air-quality monitoring. Ambient PM2.5 and ozone surveillance transformed the cardiovascular epidemiology of air pollution by providing objective, spatially resolved exposure estimates that correlate with daily MI and stroke admissions; this evidence base now underpins American Heart Association (AHA) policy statements on air-quality and cardiovascular risk.(36,71) Wastewater nicotine surveillance offers a conceptually parallel measurement, an environmental sample that integrates a community’s exposure to a major cardiovascular toxicant. Our data suggests that it too can be linked quantitatively to acute coronary events.

Prior wastewater studies of nicotine have established that cotinine, 3HC, anabasine, and anatabine can be reliably quantified in influent, that their concentrations capture temporal and spatial heterogeneity in tobacco use, that they can distinguish combustible from non-combustible products and from synthetic nicotine, and that they can be triangulated against tobacco sales records.(31,32,43–50) WBE has also been extended to a small number of chronic-disease biomarkers, including those of hypertension, diabetes, and gout in Chinese WWTPs and a wastewater signal for colorectal neoplasia in Kentucky.(72,73) However, with very few exceptions, this literature has not been examined with clinical outcome data within the same sewersheds.(74–76) Our study bridges this gap for the largest single contributor to cardiovascular mortality and, by placing the wastewater signal in geographically matched EHR data, takes WBE a step beyond descriptive exposure surveillance toward outcome-linked cardiovascular surveillance.

The observed associations are mechanistically coherent. Nicotine and its metabolites drive endothelial dysfunction, oxidative stress, vascular inflammation, accelerated atherogenesis, sympathetic activation, and platelet activation, providing a continuous pathway from chronic exposure to plaque progression and from acute exposure to thrombus initiation.(9,77–82) Within this framework, the sewershed-level associations between nicotine mass loads and the proportional burden of atherosclerosis/ischemia/infarction and heart failure mirror the chronic-exposure arm of the smoking–CVD relationship, while the selective city-level association between monthly 3HC and NMR and incident STEMI mirrors the acute-exposure arm. The fact that 3HC and the NMR (3HC/cotinine), but not cotinine, tracked STEMI is biologically informative. 3HC has a substantially shorter biological half-life than cotinine (≈6 hr vs. ≈17 hr in plasma) and, at the population level, 3HC may more sensitively reflect recent intensity of tobacco exposure rather than cumulative steady-state intake.(24,83,84) The NMR is an established phenotypic marker of CYP2A6 activity and faster nicotine clearance, which at the individual level is associated with deeper inhalation, higher daily cigarette consumption, and higher reward; the same metabolic signature may track communities with more intensive, thus more cardiotoxic tobacco use.(51–53) Consistent with this interpretation, neither biomarker was associated with NSTEMI or with chronic MI, both of which have more heterogeneous etiologies that include demand ischemia, microvascular disease, and prior remodeling rather than acute plaque rupture.(69,70) The diagnostic specificity of the wastewater signal therefore mirrors the diagnostic specificity of the underlying clinical association between active smoking and acute coronary thrombosis.

Several limitations warrant consideration. By design, wastewater nicotine concentrations serve as a proxy for aggregated smoking prevalence in the sewershed, rather than individual-level chronic exposure history. This study was not intended to capture the cumulative, years-long, pathophysiological processes underlying myocardial infarction in an individual, but rather to test whether a community-level exposure signal correlates with a corresponding community-level CVD outcome. This 12-month sampling window represents an initial feasibility assessment of wastewater-based nicotine surveillance as a novel, scalable tool for community-level CVD risk monitoring. Establishing that short-term wastewater signals correlate with acute CVD events is a necessary step toward justifying longer-term prospective designs that would be better suited to capturing the chronic exposure-outcome relationship.

As an ecological analysis, these associations are vulnerable to cross-level confounding by age, sex, socioeconomic status, ambient air pollution, diet, comorbid hypertension, diabetes, and dyslipidemia, none of which we adjusted for. The PHIX dataset was restricted to patients with at least one CVD diagnosis and did not include a non-CVD reference population. The sewershed-level analyses describe the relative composition of CVD rather than absolute population risk. The analysis is geographically restricted to a single, predominantly Hispanic city on the U.S.-Mexico border, and the small number of sewersheds (n = 4) and a 7-month overlap between wastewater and EHR datasets constrain statistical power and limit generalizability. The strength and consistency of the STEMI–3HC association across multiple analytical strategies is encouraging, but replication in larger multi-city WBE networks and over longer surveillance periods is essential before clinical or policy translation.

Although we used population-normalized mass loads to account for WWTP-specific differences in influent flow and service-area population, wastewater estimates of nicotine exposure remain subject to several quantitative uncertainties. The urinary excretion fraction of nicotine differs substantially across metabolites (approximately 10–15% as cotinine, 0.7–0.8% as nornicotine, and approximately 40% as 3HC), and in-sewer degradation, sorption to particulates, and storage stability can additionally attenuate measured signals.(23,24,85–87) We therefore report mass loads rather than reconstructed nicotine intake. Moreover, our biomarker panel does not capture all nicotine metabolites (e.g., anabasine and anatabine), and we cannot distinguish between combustible, heated-tobacco, and electronic-cigarette sources of nicotine. Future studies adding source-discriminating biomarkers will be needed to attribute the observed signal to specific tobacco product categories. All samples were collected on Mondays, which removes day-of-week heterogeneity across plants but may not be representative of average weekly exposure. Stratified random sampling across weekdays and weekends would provide a more temporally representative estimate.(88,89) Finally, factors such as the tourist and transient population flows, local events, industrial discharges, and variations within the sewer network may influence measured nicotine concentrations and were not accounted for in this analysis.

Notwithstanding these limitations, the present study positions wastewater nicotine surveillance as a candidate tool for cardiovascular population health. In contrast to retail-sales and self-reported data, wastewater integrates the entire served population in real time, captures undiagnosed and non-treatment-seeking smokers, and yields an objective, geographically resolved exposure metric whose temporal resolution is compatible with the monthly cadence of CVD surveillance. Coupled with EHR-derived incidence data, such measurements could in principle support three concrete applications relevant to the AHA mission: (i) detecting and mapping high-risk catchments to focus tobacco cessation and STEMI-preparedness resources; (ii) assessing the cardiovascular impact of tobacco-control policies (e.g., graphic warning labels, retail restrictions, indoor-air laws) by tracking the wastewater signal before and after policy changes; and (iii) monitoring the rapid evolution of the nicotine product landscape, including heated tobacco and electronic cigarettes, as it intersects with cardiovascular risk. Realizing these applications will require multi-city replication, integration with environmental and clinical covariates, harmonized analytical protocols, and prospective coupling to incident MI surveillance; given the dimensionality of CVD and tobacco-attributable risk, the potential public-health return justifies that investment.

## Author Contributions

K.M Joseph: Conceptualization, Methodology, Software, Validation, Formal Analysis, Investigations, Data Curation, Writing – Original Draft, Writing – Review & Editing, Visualization. X. Qin: Methodology, Validation, Investigation, Resources, Writing – Review & Editing. K.M. Clark: Formal Analysis, Data Curation, Writing – Review & Editing. F. Li: Methodology, Resources, Supervision, Project Administration. E. Hartmann: Resources, Data Curation. J. Nanez: Resources, Data Curation. L. Humble: Resources, Data Curation. C. Turlington: Project Administration. J. Balliew: Resources. C. Cassidy: Writing – Review & Editing. J. Narula: Funding Acquisition, Validation, Writing – Review & Editing. K.D. Mena: Conceptualization, Data Curation, Funding Acquisition, Project Administration, Supervision, Writing – Review & Editing. F. Wu: Conceptualization, Methodology, Validation, Formal Analysis, Resources, Data Curation, Writing – Original Draft, Writing – Review & Editing, Visualization, Supervision, Project Administration, Funding Acquisition.

## Data and Code Availability

All data analyzed in this study are included within the article and its supplementary materials. The analysis scripts will be made publicly available on GitHub upon publication.

## Acknowledgments

This work is supported by the National Science Foundation (DMS-2421257), the National Institute of Allergy and Infectious Diseases of the National Institutes of Health (R01AI192873), and the National Institute of Drug Abuse (R01DA059394-01A1). Additional support was provided through the K. Lance Gould Distinguished Professorship endowment (to Dr. Jagat Narula) and Dr. Craig K Cassidy is supported by the Department of Cardiology. The content is solely the responsibility of the authors and does not necessarily represent the official views of the National Institutes of Health. We acknowledge the NMR and Drug Metabolism Core at Baylor College of Medicine for the drug quantification. We are also grateful to El Paso Water for providing the samples.

## Supplementary Figures and Tables

**Figure S1.**
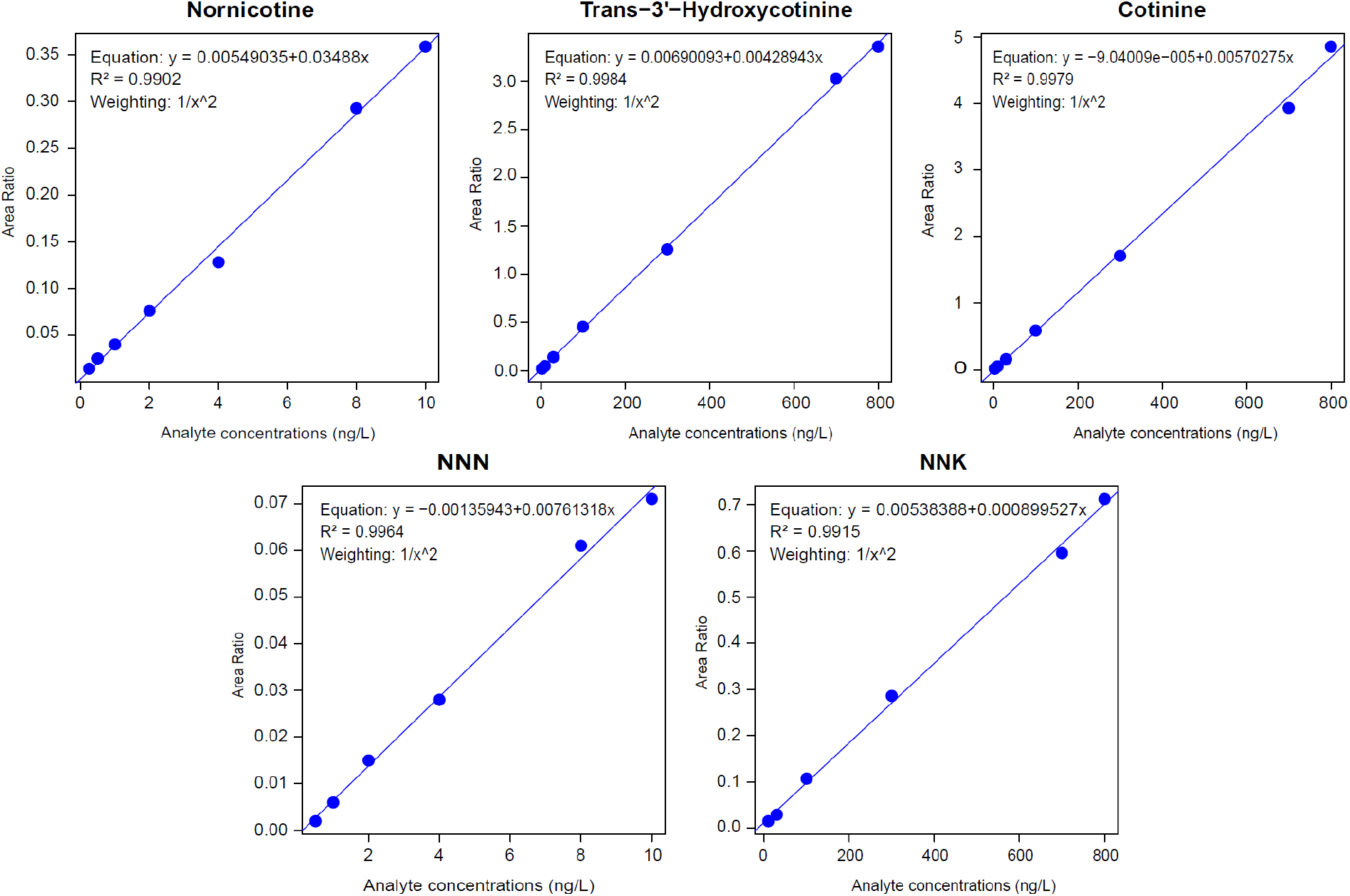
Representative standard curves for metabolite quantification of nornicotine, trans-3’-hydroxycotinine, cotinine, NNN, and NNK. Standard curves visualizing relationship between respective metabolite concentrations in water before SPE extraction (in ng/L) and peak area ratio (metabolite/IS). The analytical ranges (ng/L) and R^2^ for each metabolite include: nornicotine (0.25 – 10, 0.9902), trans-3’-hydroxycotinine (3 – 800, 0.9984), cotinine (3 – 800, 0.9979), NNN (0.5 – 10, 0.9964), and NNK (10 – 800, 0.9915). All standard curves had a weighting index of 1/X^2.

**Figure S2.**
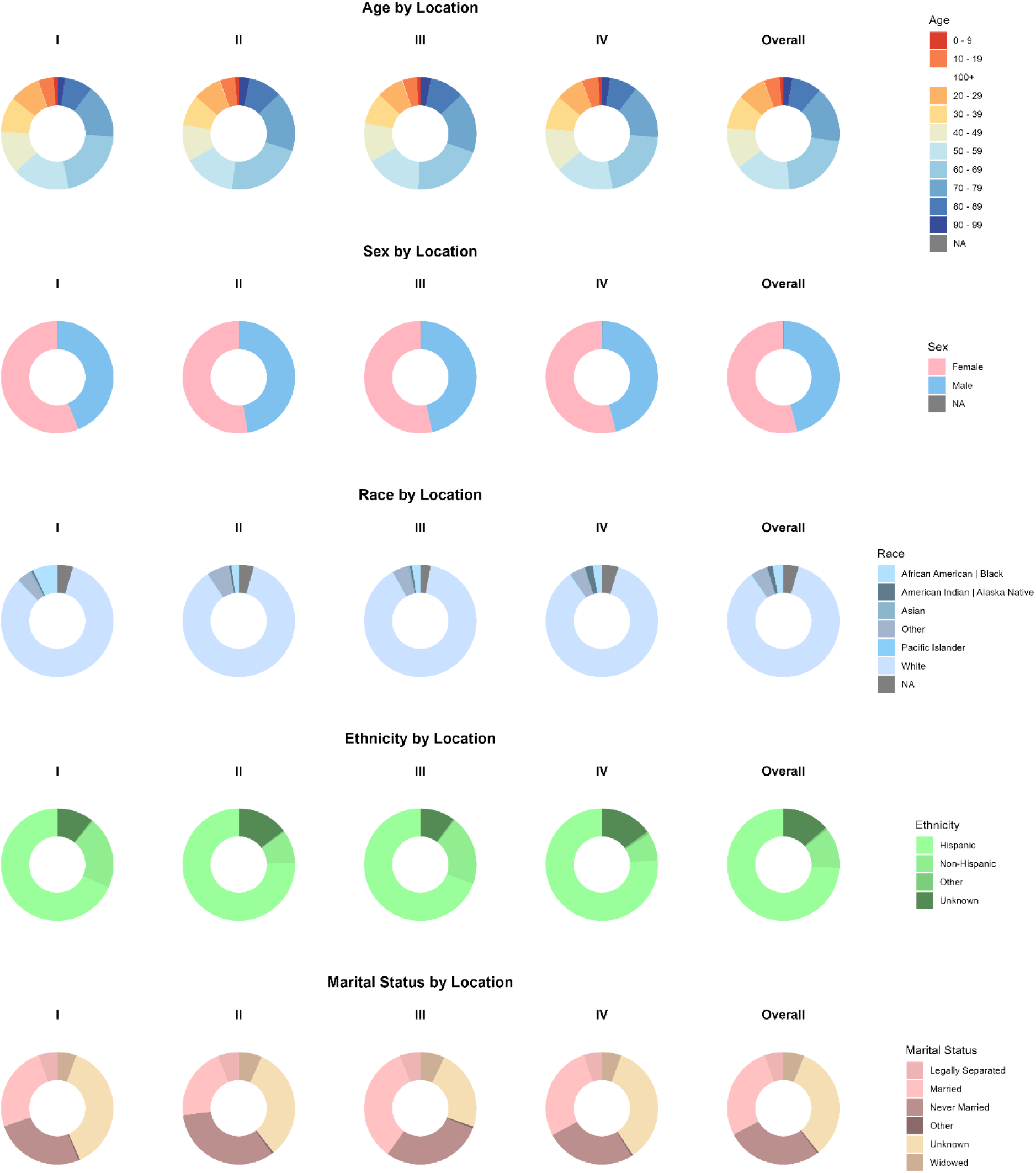
CVD Patient Demographics by WWTP. Age groups were divided by every 10 years from age 0 to over 100 years: 0-9 (1.05%), 10 – 19 (4.60%), 20-29 (8.07%), 30-39 (9.60%), 40-49 (11.93%), 50-59 (16.47%), 60-69 (20.89%), 70-79 (16.18%), 80-89 (8.53) 90-99 (2.60),100+ (0.09). Sex was classified as either Female (54.01%), Male (45.92%), or Unknown (0.06). Race was categorized by Pacific Islander (0.15%), American Indian/Alaskan Native (1.56%), Asian (0.38%), African American/Black (3.14%), White (85.87%), Other (8.89%). Ethnicity was grouped by Hispanic (73.96%), Non-Hispanic (11.83%), Other (0.60%), or Unknown (13.61%). Marital status groups were reported as Never Married (27.51%), Married (27.23%), Legally Separated (5.50%), Widowed (6.00%), Other (0.51%), Unknown (33.26%). Percentages reported here are overall for the entire patient population.

**Figure S3.**
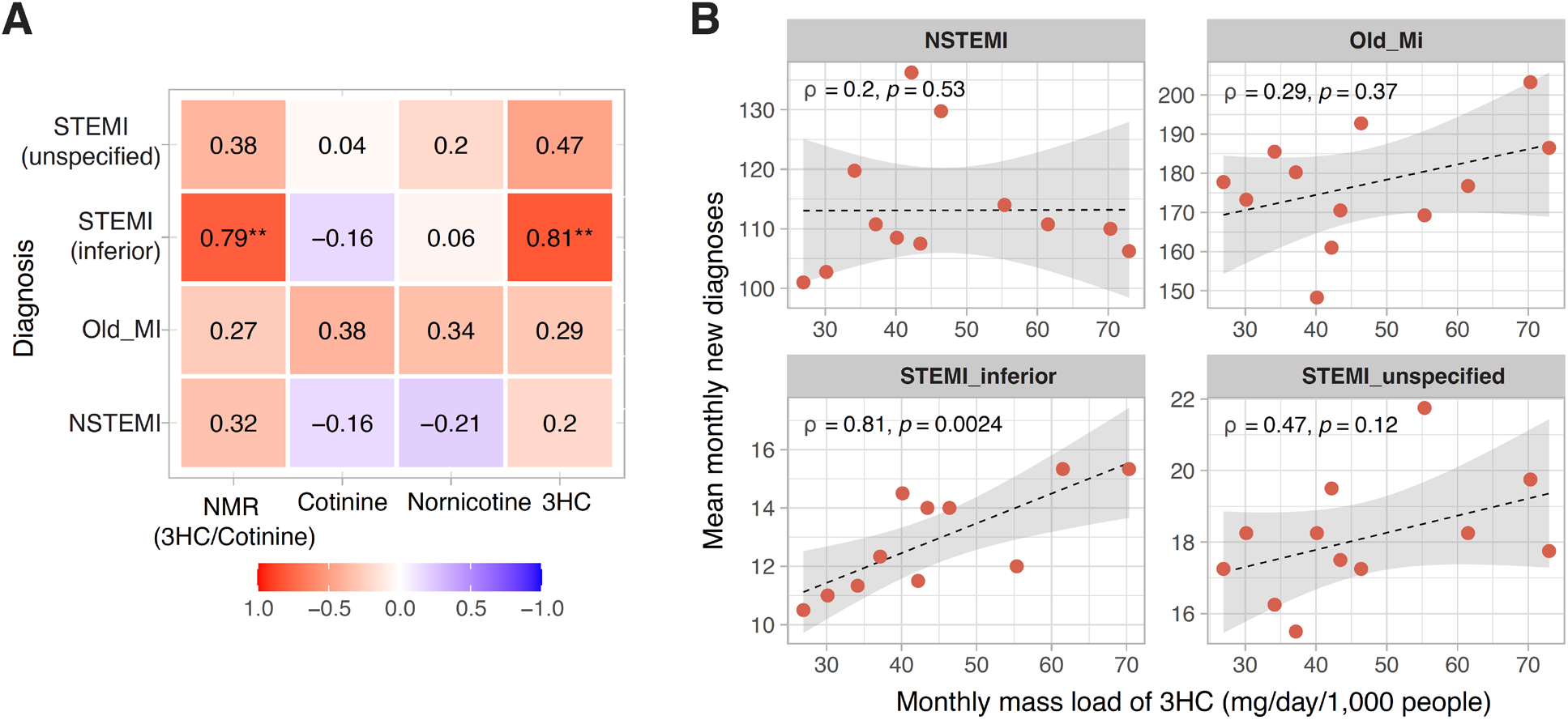
Association between monthly wastewater nicotine biomarker mass loads and mean monthly incident STEMI diagnoses at the city level using the full EHR surveillance period. (A) Heatmap of Spearman correlation coefficients (ρ) between monthly mass loads of four nicotine exposure metrics: nicotine metabolite ratio (NMR, 3HC/cotinine), cotinine, nornicotine, and 3HC, and mean monthly new diagnoses for STEMI inferior (I21.19), STEMI unspecified (I21.3), Old MI (I25.2), and NSTEMI (I21.4). CVD diagnosis counts were derived from PHIX electronic health records spanning January 2021 to December 2024, aggregated by calendar month and averaged across years to produce stable monthly estimates (n = 12 calendar months). Significance annotations reflect Spearman correlation p-values (* p < 0.05, ** p < 0.01). **(B)** Scatter plots showing the association between monthly 3HC mass load (mg/day/1,000 people) and mean monthly new diagnoses for NSTEMI, Old MI, STEMI inferior, and STEMI unspecified. Each point represents one calendar month. Dashed lines represent fitted regression lines with shaded 95% confidence intervals.

---

**Table S1.** Wastewater Flow Rates by WWTP for all samples.

| Date | Flow Rate by WWTP (in million gallons per day) |  |  |  |
| --- | --- | --- | --- | --- |
|  | I | II | III | IV |
| 6/24/2024 | 1.9 | 12.29 | 10.3 | 27.22 |
| 7/29/2024 | 7.1 | 14.53 | 8.9 | 28.31 |
| 8/26/2024 | 8.2 | 13.7 | 11.25 | 29.11 |
| 9/30/2024 | 1.8 | 9.06 | 14.94 | 27.58 |
| 10/28/2024 | 7.6 | 9.04 | 16.32 | 28.7 |
| 11/25/2024 | 2.7 | 8.99 | 19.02 | 27.3 |
| 12/30/2024 | 1.6 | 7.85 | 16.57 | 27.8 |
| 1/27/2025 | 2.8 | 8.94 | 17.03 | 27.91 |
| 2/24/2025 | 2.4 | 8.93 | 17.1 | 28.06 |
| 3/24/2025 | 7.9 | 8.6 | 18.6 | 29.16 |
| 4/28/2025 | 3.9 | 8.29 | 19.36 | 29.32 |
| 5/28/2025 | 7.4 | 5.61 | 18.45 | 29.5 |

**Table S2.** Ionization and fragmentation conditions of metabolites screened.

| Compound | Ion Transition (m/z) | Acquisition Time (min) | Collision Energy (V) | Source Fragmentation | RF Lens (V) | Retention time (min) |
| --- | --- | --- | --- | --- | --- | --- |
| Cotinine | 177→80 | 2.5-4.5 | 24.29 | 20.4 | 100 | 3.53 |
| Trans-3'-hydroxycotinine | 193→80 | 1.5-3.5 | 27.54 | 22.4 | 108 | 2.01 |
| Nornicotine | 149→132 | 0.5-2.5 | 12.96 | 0 | 72 | 1.30 |
| NNN | 178→148 | 4.0-6.0 | 10.39 | 20.4 | 63 | 4.68 |
| NNK | 208→122 | 5.0-7.0 | 13.42 | 53.1 | 154 | 5.87 |
| Cotinine-d3 (IS) | 180→80 | 2.5-4.5 | 25.35 | 22.4 | 103 | 3.52 |

**Table S3.** ICD-10 Codes of CVD conditions abstracted from PHIX, grouped by broader diagnostic categories.

| ICD-10 Code | Specific Codes Included |
| --- | --- |
| Nicotine Dependence | F17.20: Nicotine dependence, unspecified<br>• F17.200 - Nicotine dependence, unspecified, uncomplicated<br>• F17.201 – Nicotine dependence, unspecified, in remission<br>• F17.203 – Nicotine dependence unspecified, with withdrawal<br>• F17.208 – nicotine dependence, unspecified, with other nicotine-induced disorders<br>• F17.209 – nicotine dependence, unspecified, with unspecified nicotine-induced disorders<br>F17.21: Nicotine dependence, cigarettes<br>• F17.210 – Nicotine dependence, cigarettes, uncomplicated<br>• F17.211 – Nicotine dependence, cigarettes, in remission<br>• F17.213 – Nicotine dependence, cigarettes, with withdrawal<br>• F17.218 – nicotine dependence, cigarettes, with other nicotine-induced disorders<br>• F17.219 – Nicotine dependence, cigarettes, with unspecified nicotine-induced disorders<br>F17.22: Nicotine dependence, chewing tobacco<br>• F17.220 – Nicotine dependence, chewing tobacco, uncomplicated<br>• F17.221 - Nicotine dependence, chewing tobacco, in remission<br>• F17.223 - Nicotine dependence, chewing tobacco, with withdrawal<br>• F17.228 - Nicotine dependence, chewing tobacco, with other nicotine-induced disorders<br>• F17.229 - Nicotine dependence, chewing tobacco, with unspecified nicotine-induced disorders<br>F17.29: Nicotine dependence, other tobacco product |
|  | • F17.290 – Nicotine dependence, other tobacco product, uncomplicated<br>• F17.291 - Nicotine dependence, other tobacco product, in remission<br>• F17.293 - Nicotine dependence, other tobacco product, with withdrawal<br>• F17.298 - Nicotine dependence, other tobacco product, with other nicotine-induced disorders<br>• F17.299 - Nicotine dependence, other tobacco product, with unspecified nicotine-induced disorders |
| Abnormal Heart Rhythm | Abnormalities of Heart Rhythm (ICD-9-CM 427.81, 427.89, 785.0, 785.1, 785.3)<br>• R00.0 – tachycardia, unspecified<br>• R00.1 – bradycardia, unspecified<br>• R00.2 – palpitations<br>• R00.8 – other abnormalities of heart beat<br>• R00.9 – unspecified abnormalities of heart beat<br>Atrial fibrillation and flutter (ICD-9-CM 427.31, 427.32)<br>• I48.0 – paroxysmal atrial fibrillation<br>• I48.1 – persistent atrial fibrillation<br>• I48.2 – chronic atrial fibrillation<br>• I48.3 – typical atrial flutter<br>• I48.4 – atypical atrial flutter<br>• I48.91 – unspecified atrial fibrillation<br>• I48.92 – unspecified atrial flutter<br>Cardiac Arrhythmias (Other) (ICD-9-CM 427.41, 427.42, 427.60, 427.61, 427.69, 427.81, 427.89, 427.9)<br>• I49.01 – ventricular fibrillation<br>• I49.02 – ventricular flutter<br>• I49.1 - Atrial premature depolarization<br>• I49.2 - Junctional premature depolarization<br>• I49.3 - Ventricular premature depolarization<br>• I49.40 – unspecified premature depolarization<br>• I49.49 – other premature depolarization<br>• I49.5 – sick sinus syndrome |
|  | • I49.8 – other specified cardiac arrhythmias<br>• I49.9 – cardiac arrhythmia, unspecified |
| Chest Pain | Chest Pain (ICD-9-CM 411.1, 413.1, 413.9, 786.50 – 786.59)<br>• I20.0 – unstable angina<br>• I20.1 – angina pectoris with documented spasm<br>• I20.8 – other forms of angina pectoris<br>• I20.9 – angina pectoris, unspecified<br>• R07.1 – chest pain on breathing<br>• R07.2 – precordial pain<br>• R07.81 – pleurodynia<br>• R07.82 – intercostal pain<br>• R07.89 – other chest pain<br>• R07.9 – chest pain, unspecified |
| Heart Failure | Heart failure (ICD-9-CM 428.0, 428.1, 428.20 to 428.23 range, 428.30 to 428.33 range, 428.40 – 428.43 range, 428.9)<br>• I50.1 – left ventricular failure<br>• I50.20 – unspecified systolic (congestive) heart failure<br>• I50.21 – acute systolic (congestive) heart failure<br>• I50.22 – chronic systolic (congestive) heart failure<br>• I50.23 – acute on chronic systolic (congestive) heart failure<br>• I50.30 – unspecified diastolic (congestive) heart failure<br>• I50.31 – acute diastolic (congestive) heart failure<br>• I50.32 – chronic diastolic (congestive) heart failure<br>• I50.33 – acute on chronic diastolic (congestive) heart failure<br>• I50.40 – unspecified combined systolic (congestive) and diastolic (congestive) heart failure<br>• I50.41 – acute combined systolic (congestive) and diastolic (congestive) heart failure<br>• I50.42 – chronic combined systolic (congestive) and diastolic (congestive) heart failure<br>• I50.43 – acute on chronic combined systolic (congestive) and diastolic (congestive) heart failure |
|  | • I50.9 – heart failure, unspecified |
| Hypertension | Hypertension (ICD-9-CM 401.9)<br>• I10 – essential (primary) hypertension |
| Nonrheumatic valve disorders | Nonrheumatic valve disorders – aortic valve disorders (ICD-9-CM 424.1)<br>• I35.0 – nonrheumatic aortic (valve) stenosis<br>• I35.1 – nonrheumatic aortic (valve) insufficiency<br>• I35.2 – nonrheumatic aortic (valve) stenosis with insufficiency<br>• I35.8 – other nonrheumatic aortic valve stenosis<br>• I35.9 – nonrheumatic aortic valve disorder, unspecified<br>Nonrheumatic valve disorders – mitral valve disorders (ICD-9-CM 424.0)<br>• I34.0 – nonrheumatic mitral (valve) insufficiency<br>• I34.1 – nonrheumatic mitral (valve) prolapse<br>• I34.2 – nonrheumatic mitral (valve) stenosis<br>• I34.8 – other nonrheumatic mitral valve insufficiency<br>• I34.9 – nonrheumatic mitral valve disorder, unspecified |
| Atherosclerosis | Selected Atherosclerosis, Ischemia, and Infarction (ICD-9-CM 410.00 to 410.92 Range, 411.1, 412, 413.0, 413.1, 413.9, 414.00 to 414.07 Range, 414.10, 414.11, 414.12, 414.19, 414.2, 414.3, 414.4, 414.8, 414.9, 429.2, 429.5, 429.6, 429.71, 429.79)<br>• I21.01 – ST elevation (STEMI) myocardial infarction involving left main coronary artery<br>• I21.02 – ST elevation (STEMI) myocardial infarction involving left anterior descending coronary artery<br>• I21.09 - ST elevation (STEMI) myocardial infarction involving other coronary artery of anterior wall<br>• I21.11 - ST elevation (STEMI) myocardial infarction involving right coronary artery<br>• I21.19 - ST elevation (STEMI) myocardial infarction involving other coronary artery of interior wall<br>• I21.21 - ST elevation (STEMI) myocardial infarction involving left circumflex coronary artery<br>• I21.29 - ST elevation (STEMI) myocardial infarction involving other sites |
|  | • I21.3 - ST elevation (STEMI) myocardial infarction of unspecified sites<br>• I21.4 – Non-ST elevation (NSTEMI) myocardial infarction<br>• I22.0 – Subsequent ST elevation (STEMI) myocardial infarction of anterior wall<br>• I22.1 – Subsequent ST elevation (STEMI) myocardial infarction of inferior wall<br>• I22.2 – Subsequent non-ST elevation (NSTEMI) myocardial infarction<br>• I22.8 – Subsequent ST elevation (STEMI) myocardial infarction of other sites<br>• I22.9 – subsequent ST elevation (STEMI) myocardial infarction of unspecified site<br>• I23.0 – Hemopericardium as current complication following acute myocardial infarction<br>• I23.1 – Atrial septal defect as current complication following acute myocardial infarction<br>• I23.2 – Ventricular septal defect as current complication following acute myocardial infarction<br>• I23.3 – Rupture of cardiac wall without hemopericardium as current complication following acute myocardial infarction<br>• I23.4 – Rupture of chordae tendineae as current complication following acute myocardial infarction<br>• I23.5 – rupture of papillary muscle as current complication following acute myocardial infarction<br>• I23.6 – thrombosis of atrium, auricular appendage, and ventricle as current complications following acute myocardial infarction<br>• I23.7 – postinfarction angina<br>• I23.8 – other current complications following acute myocardial infarction<br>• 125.10 – Atherosclerotic heart disease of native coronary artery without angina pectoris<br>• 125.110 – Atherosclerotic heart disease of native coronary artery with unstable angina pectoris<br>• 125.111 – Atherosclerotic heart disease of native coronary artery with angina pectoris with documented muscle spasm |
|  | • 125.118 – Atherosclerotic heart disease of native coronary artery with other forms of angina pectoris<br>• 125.119 – Atherosclerotic heart disease of native coronary artery with unspecified angina pectoris<br>• I25.2 – old myocardial infarction<br>• I25.3 – aneurysm of heart<br>• I25.41 – coronary artery aneurysm<br>• I25.42 – coronary artery dissection<br>• I25.5 – ischemic cardiomyopathy<br>• I25.6 – silent myocardial ischemia<br>• I25.700 – atherosclerosis of coronary artery bypass graft(s), unspecified, with unstable angina pectoris<br>• I25.701 - atherosclerosis of coronary artery bypass graft(s), unspecified, with angina pectoris with documented spasm<br>• I25.708 - atherosclerosis of coronary artery bypass graft(s), unspecified, with other forms of angina pectoris<br>• I25.709 - atherosclerosis of coronary artery bypass graft(s), unspecified, with unspecified angina pectoris<br>• I25.710 - atherosclerosis of autologous veing coronary artery bypass graft(s) with unstable angina pectoris<br>• I25.711 - atherosclerosis of autologous vein coronary artery bypass graft(s) with angina pectoris with documented spasm<br>• I25.718 - atherosclerosis of autologous vein coronary artery bypass graft(s) with other forms of angina pectoris<br>• I25.719 - atherosclerosis of autologous vein coronary artery bypass graft(s) with unspecified forms of angina pectoris<br>• I25.720 - atherosclerosis of autologous artery coronary artery bypass graft(s) with unstable angina pectoris<br>• I25.721 – atherosclerosis of autologous artery coronary artery bypass graft(s) with angina pectoris with documented spasm<br>• I25.728 – Atherosclerosis of autologous artery coronary artery bypass graft(s) with other forms of angina pectoris<br>• I25.729 – atherosclerosis of autologous artery coronary artery bypass graft(s) with unspecified angina pectoris |
|  | • I25.730 – atherosclerosis of nonautologous biological coronary artery bypass graft(s) with unstable angina pectoris<br>• I25.731 – atherosclerosis of nonautologous biological coronary artery bypass graft(s) with angina pectoris with documented spasm<br>• I25.738 – atherosclerosis of nonautologous biological coronary artery bypass graft(s) with other forms of angina pectoris<br>• I25.739 - atherosclerosis of nonautologous biological coronary artery bypass graft(s) with unspecified angina pectoris<br>• I25.750 – atherosclerosis of native coronary artery of transplanted heart with unstable angina<br>• I25.751 - atherosclerosis of native coronary artery of transplanted heart with angina pectoris with documented spasm<br>• I25.758 – atherosclerosis of native coronary artery of transplanted heart with other forms of angina pectoris<br>• I25.759 – atherosclerosis of native coronary artery of transplanted heart with unspecified angina pectoris<br>• I25.760 – atherosclerosis of bypass graft of coronary artery of transplanted heart with unstable angina<br>• I25.761 – atherosclerosis of bypass graft of coronary artery of transplanted heart with angina pectoris with documented spasm<br>• I25.768 – atherosclerosis of bypass graft of coronary artery of transplanted heart with other forms of angina pectoris<br>• I25.769 – atherosclerosis of bypass graft of coronary artery of transplanted heart with unspecified angina pectoris<br>• I25.790 – atherosclerosis of other coronary artery bypass graft(s) with unstable angina pectoris<br>• I25.791 – atherosclerosis of other coronary artery bypass graft(s) with angina pectoris with documented spasm<br>• I25.798 – atherosclerosis of other coronary artery bypass graft(s) with other forms of angina pectoris<br>• I25.799 – atherosclerosis of other coronary artery bypass graft(s) with unspecified angina pectoris |
|  | • I25.810 – atherosclerosis of coronary artery bypass graft(s) without angina pectoris<br>• I25.811 – atherosclerosis of native coronary artery of transplanted heart without angina pectoris<br>• I25.812 – atherosclerosis of bypass graft of coronary artery of transplanted heart without angina pectoris<br>• I25.82 – chronic total occlusion of coronary artery<br>• I25.83 – coronary atherosclerosis due to lipid rich plaque<br>• I25.84 – coronary atherosclerosis due to calcified coronary lesion<br>• I25.89 – other forms of chronic ischemic heart disease<br>• I25.9 – chronic ischemic heart disease, unspecified |
| Syncope and Collapse | Syncope and Collapse<br>R55 – syncope and collapse |
| Unstable Angina | • I20.0 – unstable angina |
| Old Myocardial Infarction | • I25.2 – old myocardial infarction |
| NSTEMI (I21.4) | • I21.4 – Non-ST elevation (NSTEMI) myocardial infarction |
| STEMI (I21.09) | • I21.09 - ST elevation (STEMI) myocardial infarction involving other coronary artery of anterior wall |
| STEMI (I21.19) | • I21.19 - ST elevation (STEMI) myocardial infarction involving other coronary artery of interior wall |
| STEMI (I21.29) | • I21.29 - ST elevation (STEMI) myocardial infarction involving other sites |
| STEMI (I21.3) | • I21.3 - ST elevation (STEMI) myocardial infarction of unspecified sites |
| STEMI (I22.9) | • I22.9 – subsequent ST elevation (STEMI) myocardial infarction of unspecified site |

## References

1 Achievements in Public Health, 1900-1999: Decline in Deaths from Heart Disease and Stroke -- United States, 1900-1999 [Internet]. [cited 2026 Jan 1]. Available from: https://www.cdc.gov/mmwr/preview/mmwrhtml/mm4830a1.htm

2. Dalen JE, Alpert JS, Goldberg RJ, Weinstein RS. The Epidemic of the 20th Century: Coronary Heart Disease. The American Journal of Medicine. 2014 Sep 1;127(9):807–12. doi:10.1016/j.amjmed.2014.04.015 PubMed PMID: 24811552.

3. Haidar A, Gajjar A, Parikh RV, Benharash P, Fonarow GC, Watson K, et al. National Costs for Cardiovascular-Related Hospitalizations and Inpatient Procedures in the United States, 2016 to 2021. American Journal of Cardiology. 2025 Jan 1;234:63–70. doi:10.1016/j.amjcard.2024.10.003 PubMed PMID: 39454699.

4. Cardiovascular diseases (CVDs) [Internet]. [cited 2026 Jan 6]. Available from: https://www.who.int/news-room/fact-sheets/detail/cardiovascular-diseases-(cvds)

5. Olvera Lopez E, Ballard BD, Jan A. Cardiovascular Disease. In: StatPearls [Internet]. Treasure Island (FL): StatPearls Publishing; 2025 [cited 2026 Jan 1]. Available from: http://www.ncbi.nlm.nih.gov/books/NBK535419/ PubMed PMID: 30571040.

6. Mayo Clinic [Internet]. [cited 2026 Jan 6]. Heart disease - Symptoms and causes. Available from: https://www.mayoclinic.org/diseases-conditions/heart-disease/symptoms-causes/syc-20353118

7. Definition of cardiovascular disease - NCI Dictionary of Cancer Terms - NCI [nciAppModulePage] [Internet]. 2011 [cited 2026 Jan 6]. Located at: nciglobal,ncienterprise. Available from: https://www.cancer.gov/publications/dictionaries/cancer-terms/def/cardiovascular-disease

8. Mitsis A, Gragnano F. Myocardial Infarction with and without ST-segment Elevation: a Contemporary Reappraisal of Similarities and Differences. Curr Cardiol Rev. 2021 Jul 20;17(4):e230421189013. doi:10.2174/1573403X16999201210195702 PubMed PMID: 33305709; PubMed Central PMCID: PMC8762150.

9. Alhindal M, Janahi J, D’Angelo EC, Lisignoli V, Palmieri R, Cutrì A, et al. Impact of smoking on cardiovascular health: Mechanisms, epidemiology and specific concerns regarding congenital heart disease. International Journal of Cardiology Congenital Heart Disease. 2025 Jun 1;20:100581. doi:10.1016/j.ijcchd.2025.100581

10. Gallucci G, Tartarone A, Lerose R, Lalinga AV, Capobianco AM. Cardiovascular risk of smoking and benefits of smoking cessation. J Thorac Dis. 2020 Jul;12(7):3866–76. doi:10.21037/jtd.2020.02.47 PubMed PMID: 32802468; PubMed Central PMCID: PMC7399440.

11 Definition of free radical - NCI Dictionary of Cancer Terms - NCI [nciAppModulePage] [Internet]. 2011 [cited 2026 Jan 1]. Located at: nciglobal,ncienterprise. Available from: https://www.cancer.gov/publications/dictionaries/cancer-terms/def/free-radical

12. Lee J, Taneja V, Vassallo R. Cigarette Smoking and Inflammation. J Dent Res. 2012 Feb;91(2):142–9. doi:10.1177/0022034511421200 PubMed PMID: 21876032; PubMed Central PMCID: PMC3261116.

13. Rahman M, Alatiqi M, Al Jarallah M, Hussain MY, Monayem A, Panduranga P, et al. Cardiovascular Effects of Smoking and Smoking Cessation: A 2024 Update. Glob Heart. 20(1):15. doi:10.5334/gh.1399 PubMed PMID: 39991592; PubMed Central PMCID: PMC11843939.

14. Dennison Himmelfarb CR, Benowitz NL, Blank MD, Bhatnagar A, Chase PJ, Davis EM, et al. Impact of Smokeless Oral Nicotine Products on Cardiovascular Disease: Implications for Policy, Prevention, and Treatment: A Policy Statement From the American Heart Association. Circulation. 2025 Jan 7;151(1):e1–21. doi:10.1161/CIR.0000000000001293

15. O’Connor R, Schneller LM, Felicione NJ, Talhout R, Goniewicz ML, Ashley DL. Evolution of tobacco products: recent history and future directions. Tobacco Control. 2022 Mar 1;31(2):175–82. doi:10.1136/tobaccocontrol-2021-056544 PubMed PMID: 35241585.

16. Lichtenberg K. E-Cigarettes: Current Evidence and Policy. Mo Med. 2017;114(5):335–8. PubMed PMID: 30228625; PubMed Central PMCID: PMC6140188.

17. Espinoza-Derout J, Shao XM, Lao CJ, Hasan KM, Rivera JC, Jordan MC, et al. Electronic Cigarette Use and the Risk of Cardiovascular Diseases. Front Cardiovasc Med. 2022 Apr 7;9:879726. doi:10.3389/fcvm.2022.879726 PubMed PMID: 35463745; PubMed Central PMCID: PMC9021536.

18. Zong H, Hu Z, Li W, Wang M, Zhou Q, Li X, et al. Electronic cigarettes and cardiovascular disease: epidemiological and biological links. Pflugers Arch - Eur J Physiol. 2024 Jun 1;476(6):875–88. doi:10.1007/s00424-024-02925-0

19. Chaiton M, Pienkowski M, Musani I, Bondy SJ, Cohen JE, Dubray J, et al. Smoking, e-cigarettes and the effect on respiratory symptoms among a population sample of youth: Retrospective cohort study. Tob Induc Dis. 2023 Jan 21;21:08. doi:10.18332/tid/156839 PubMed PMID: 36721859; PubMed Central PMCID: PMC9865633.

20. Matthews J, Matthews M, Cherian V. A cloud of addiction: how vaping has created a new generation of addicts. Br J Gen Pract. 2023 Jul 1;73(suppl 1). doi:10.3399/bjgp23X734325 PubMed PMID: 37479276.

21. Magna A, Polisena N, Polisena L, Bagnato C, Pacella E, Carnevale R, et al. The Hidden Dangers: E-Cigarettes, Heated Tobacco, and Their Impact on Oxidative Stress and Atherosclerosis—A Systematic Review and Narrative Synthesis of the Evidence. Antioxidants (Basel). 2024 Nov 15;13(11):1395. doi:10.3390/antiox13111395 PubMed PMID: 39594537; PubMed Central PMCID: PMC11591068.

22. Wold LE, Tarran R, Crotty Alexander LE, Hamburg NM, Kheradmand F, St. Helen G, et al. Cardiopulmonary Consequences of Vaping in Adolescents: A Scientific Statement From the American Heart Association. Circulation Research. 2022 Jul 22;131(3):e70–82. doi:10.1161/RES.0000000000000544

23. Benowitz NL, Hukkanen J, Jacob P. Nicotine Chemistry, Metabolism, Kinetics and Biomarkers. Handb Exp Pharmacol. 2009;(192):29–60. doi:10.1007/978-3-540-69248-5_2 PubMed PMID: 19184645; PubMed Central PMCID: PMC2953858.

24. Benowitz NL, Jacob P. Trans-3′-hydroxycotinine: Disposition kinetics, effects and plasma levels during cigarette smoking. Br J Clin Pharmacol. 2001 Jan;51(1):53–9. doi:10.1046/j.1365-2125.2001.01309.x PubMed PMID: 11167665; PubMed Central PMCID: PMC2014428.

25. Siminszky B, Gavilano L, Bowen SW, Dewey RE. Conversion of nicotine to nornicotine in Nicotiana tabacum is mediated by CYP82E4, a cytochrome P450 monooxygenase. Proc Natl Acad Sci U S A. 2005 Oct 11;102(41):14919–24. doi:10.1073/pnas.0506581102 PubMed PMID: 16192354; PubMed Central PMCID: PMC1253577.

26. Hoffmann D, Hecht SS, Ornaf RM, Wynder EL. N′-Nitrosonornicotine in Tobacco. Science. 1974 Oct 18;186(4160):265–7. doi:10.1126/science.186.4160.265

27. Stepanov I, Sebero E, Wang R, Gao YT, Hecht SS, Yuan JM. Tobacco-specific N-nitrosamine exposures and cancer risk in the Shanghai Cohort Study: remarkable coherence with rat tumor sites. Int J Cancer. 2014 May 15;134(10):2278–83. doi:10.1002/ijc.28575 PubMed PMID: 24243522; PubMed Central PMCID: PMC3949147.

28. Maertens LA, Upadhyaya P, Hecht SS, Zimmerman CL. Formation and Distribution of NNK Metabolites in an Isolated Perfused Rat Lung. Drug Metab Dispos. 2010 May;38(5):752–60. doi:10.1124/dmd.109.031492 PubMed PMID: 20159989; PubMed Central PMCID: PMC2872947.

29. Wang SQ, Bao LJ, Li TY, Zeng EY. Potential health risk of human exposure to tobacco-specific nitrosamines in second-hand and third-hand smoke. Journal of Hazardous Materials. 2024 Dec 5;480:136446. doi:10.1016/j.jhazmat.2024.136446

30. Schick SF, Blount BC, Jacob P, Saliba NA, Bernert JT, El Hellani A, et al. Biomarkers of exposure to new and emerging tobacco delivery products. Am J Physiol Lung Cell Mol Physiol. 2017 Sep 1;313(3):L425–52. doi:10.1152/ajplung.00343.2016 PubMed PMID: 28522563; PubMed Central PMCID: PMC5626373.

31. Asadi A, Fakhri Y, Salimi Y, Daglioglu N, Tahmasebifard M, Aghajarinezhad M. Nicotine consumption rate through wastewater-based epidemiology: a systematic review, meta-analysis and probabilistic risk assessment. Environ Sci Pollut Res Int. 2023;30(23):63416–26. doi:10.1007/s11356-023-27017-x PubMed PMID: 37084052; PubMed Central PMCID: PMC10119841.

32. Weng MT, Steadman KJ, Zhong K, Al-Gharibeh M, Zheng Q, Thai PK. Difference in nicotine metabolism as a potential factor causing disparities in tobacco consumption estimates by wastewater-based epidemiology. Health Sciences Review. 2025 Mar 1;14:100210. doi:10.1016/j.hsr.2025.100210

33. Diamond MB, Keshaviah A, Bento AI, Conroy-Ben O, Driver EM, Ensor KB, et al. Wastewater surveillance of pathogens can inform public health responses. Nat Med. 2022 Oct;28(10):1992–5. doi:10.1038/s41591-022-01940-x

34. National Academies of Sciences E, Division H and M, Studies D on E and L, Practice B on PH and PH, Board WS and T, Surveillance C on CW based ID. Introduction. In: Wastewater-based Disease Surveillance for Public Health Action [Internet]. National Academies Press (US); 2023 [cited 2026 May 5]. Available from: https://www.ncbi.nlm.nih.gov/books/NBK591714/

35. National Academies of Sciences E, Division H and M, Studies D on E and L, Practice B on PH and PH, Board WS and T, Surveillance C on CW based ID. Vision for National Wastewater Surveillance. In: Wastewater-based Disease Surveillance for Public Health Action [Internet]. National Academies Press (US); 2023 [cited 2026 May 5]. Available from: https://www.ncbi.nlm.nih.gov/books/NBK591717/

36. Olaniyan T, Pinault L, Li C, van Donkelaar A, Meng J, Martin RV, et al. Ambient air pollution and the risk of acute myocardial infarction and stroke: A national cohort study. Environmental Research. 2022 Mar 1;204:111975. doi:10.1016/j.envres.2021.111975

37. Impact of Short-Term Exposure to Ozone on Hospital Admissions for Multiple Cardiovascular Diseases: A Systematic Review and Meta-Analysis | Journal of the American Heart Association [Internet]. [cited 2026 May 14]. Available from: https://www.ahajournals.org/doi/10.1161/JAHA.124.037205

38. Miller KA, Siscovick DS, Sheppard L, Shepherd K, Sullivan JH, Anderson GL, et al. Long-Term Exposure to Air Pollution and Incidence of Cardiovascular Events in Women. New England Journal of Medicine. 2007 Feb 1;356(5):447–58. doi:10.1056/NEJMoa054409

39. Chen X, Balliew J, Bauer CX, Deegan J, Gitter A, Hanson BM, et al. Revealing patterns of SARS-CoV-2 variant emergence and evolution using RBD amplicon sequencing of wastewater. Journal of Infection. 2024 Nov 1;89(5). doi:10.1016/j.jinf.2024.106284 PubMed PMID: 39341403.

40. Oghuan J, Chavarria C, Vanderwal SR, Gitter A, Ojaruega AA, Monserrat C, et al. Wastewater analysis of Mpox virus in a city with low prevalence of Mpox disease: an environmental surveillance study. Lancet Reg Health Am. 2023 Nov 24;28:100639. doi:10.1016/j.lana.2023.100639 PubMed PMID: 38076410; PubMed Central PMCID: PMC10701415.

41. Barcellos DS, Barquilha CER, Oliveira PE, Prokopiuk M, Etchepare RG. How has the COVID-19 pandemic impacted wastewater-based epidemiology? Sci Total Environ. 2023 Sep 20;892:164561. doi:10.1016/j.scitotenv.2023.164561 PubMed PMID: 37263440; PubMed Central PMCID: PMC10226898.

42. Gitter A, Oghuan J, Godbole AR, Chavarria CA, Monserrat C, Hu T, et al. Not a waste: Wastewater surveillance to enhance public health. Front Chem Eng. 2023 Jan 9;4:1112876. doi:10.3389/fceng.2022.1112876

43. Zheng Q, Gartner C, Tscharke BJ, O’Brien JW, Gao J, Ahmed F, et al. Long-term trends in tobacco use assessed by wastewater-based epidemiology and its relationship with consumption of nicotine containing products. Environ Int. 2020 Dec;145:106088. doi:10.1016/j.envint.2020.106088 PubMed PMID: 32911244.

44. Zheng Q, Gerber C, Steadman KJ, Lin CY, Tscharke BJ, O’Brien JW, et al. Improving Wastewater-Based Tobacco Use Estimates Using Anabasine. Environ Sci Technol. 2023 May 30;57(21):7958–65. doi:10.1021/acs.est.3c01510

45. Wang Z, Zheng Q, O’Brien JW, Tscharke BJ, Chan G, Thomas KV, et al. Analysis of wastewater from 2013 to 2021 detected a recent increase in nicotine use in Queensland, Australia. Water Research. 2024 Feb 15;250:121040. doi:10.1016/j.watres.2023.121040

46. Zheng Q, Eaglesham G, Tscharke BJ, O’Brien JW, Li J, Thompson J, et al. Determination of anabasine, anatabine, and nicotine biomarkers in wastewater by enhanced direct injection LC-MS/MS and evaluation of their in-sewer stability. Science of The Total Environment. 2020 Nov 15;743:140551. doi:10.1016/j.scitotenv.2020.140551

47. Tscharke BJ, White JM, Gerber JP. Estimates of tobacco use by wastewater analysis of anabasine and anatabine. Drug Testing and Analysis. 2016;8(7):702–7. doi:10.1002/dta.1842

48. Lai FY, Gartner C, Hall W, Carter S, O’Brien J, Tscharke BJ, et al. Measuring spatial and temporal trends of nicotine and alcohol consumption in Australia using wastewater-based epidemiology. Addiction. 2018;113(6):1127–36. doi:10.1111/add.14157

49. Cheetham AG, Plunkett S, Campbell P, Hilldrup J, Coffa BG, Gilliland S, et al. Analysis and differentiation of tobacco-derived and synthetic nicotine products: Addressing an urgent regulatory issue. PLoS One. 2022 Apr 14;17(4):e0267049. doi:10.1371/journal.pone.0267049 PubMed PMID: 35421170; PubMed Central PMCID: PMC9009602.

50. Head C, Chanakira E, Gillespie D, Breeze P, Angus C, Thomas C, et al. Wastewater-based epidemiology for local tobacco and nicotine use with small-area estimations. Eur J Public Health. 2025 Oct 27;35(Suppl 4):ckaf161.1141. doi:10.1093/eurpub/ckaf161.1141 PubMed PMID: null; PubMed Central PMCID: PMC12555257.

51. Gubner NR, Guydish J, Humfleet GL, Benowitz NL, Hall SM. Nicotine biomarkers and rate of nicotine metabolism among cigarette smokers taking buprenorphine for opioid dependency. Drug Alcohol Depend. 2017 Sep 1;178:267–70. doi:10.1016/j.drugalcdep.2017.05.020 PubMed PMID: 28683421; PubMed Central PMCID: PMC5553616.

52. Buchwald J, Chenoweth MJ, Palviainen T, Zhu G, Benner C, Gordon S, et al. Genome-wide association meta-analysis of nicotine metabolism and cigarette consumption measures in smokers of European descent. Mol Psychiatry. 2021 Jun;26(6):2212–23. doi:10.1038/s41380-020-0702-z PubMed PMID: 32157176; PubMed Central PMCID: PMC7483250.

53. Sofuoglu M, Herman AI, Nadim H, Jatlow P. Rapid Nicotine Clearance is Associated with Greater Reward and Heart Rate Increases from Intravenous Nicotine. Neuropsychopharmacology. 2012 May;37(6):1509–16. doi:10.1038/npp.2011.336 PubMed PMID: 22334123; PubMed Central PMCID: PMC3327855.

54. Kuang Z, Lin L, Kong R, Wang Z, Mao X, Xiang D. The correlation between cumulative cigarette consumption and infarction-related coronary spasm in patients with ST-segment elevation acute myocardial infarction across different age groups. Sci Rep. 2025 Jan 2;15(1):253. doi:10.1038/s41598-024-84125-5

55. Palmer J, Lloyd A, Steele L, Fotheringham J, Teare D, Iqbal J, et al. Differential Risk of ST-Segment Elevation Myocardial Infarction in Male and Female Smokers. JACC. 2019 Jul 2;73(25):3259–66. doi:10.1016/j.jacc.2019.03.525

56. Boulos M, Sharif Y, Assy N, Sharif D. Significance of Smoking in Patients with Acute ST Elevation Myocardial Infarction (STEMI) Undergoing Primary Percutaneous Coronary Intervention: Evaluation of Coronary Flow, Microcirculation and Left Ventricular Systolic Function. Hearts. 2024 Mar;5(1):182–95. doi:10.3390/hearts5010012

57. US judge blocks FDA graphic warning label requirement for cigarettes | Reuters [Internet]. [cited 2026 Jan 6]. Available from: https://www.reuters.com/legal/us-judge-blocks-fda-graphic-warning-label-requirement-cigarettes-2025-01-14/

58. Devi S. Texan judge blocks US FDA changes on cigarette packaging. The Lancet Oncology. 2025 Feb 1;26(2):e79. doi:10.1016/S1470-2045(25)00023-3 PubMed PMID: 39864446.

59. Joseph KM, Qin X, Parikh D, Li F, Balliew J, Mena KD, et al. First Detection of Xylazine in Texas Wastewater and Its Association with Fentanyl Use. ACS EST Water. 2026 Feb 13;6(2):946–53. doi:10.1021/acsestwater.5c01016

60. Food and Drug Administration. Bioanalytical Method Validation Guidance for Industry [Internet]. US Food and Drug Administration; 2018 [cited 2025 Aug 7]. Located at: FDA-2013-D-1020. Available from: https://www.fda.gov/regulatory-information/search-fda-guidance-documents/bioanalytical-method-validation-guidance-industry

61. Fontanals N, Marcé RM, Montes R, Rodil R, González-Mariño I, Valcárcel Y, et al. Wastewater-based epidemiology to assess pharmaceutical consumption. Spanish perspective. Science of The Total Environment. 2024 Nov 25;953:176108. doi:10.1016/j.scitotenv.2024.176108

62. Bishop N, Jones-Lepp T, Margetts M, Sykes J, Alvarez D, Keil DE. Wastewater-based epidemiology pilot study to examine drug use in the Western United States. Sci Total Environ. 2020 Nov 25;745:140697. doi:10.1016/j.scitotenv.2020.140697 PubMed PMID: 32758738; PubMed Central PMCID: PMC7361084.

63. Gushgari AJ, Venkatesan AK, Chen J, Steele JC, Halden RU. Long-term tracking of opioid consumption in two United States cities using wastewater-based epidemiology approach. Water Res. 2019 Sep 15;161:171–80. doi:10.1016/j.watres.2019.06.003 PubMed PMID: 31195333; PubMed Central PMCID: PMC6613989.

64. Castiglioni S, Bijlsma L, Covaci A, Emke E, Hernández F, Reid M, et al. Evaluation of Uncertainties Associated with the Determination of Community Drug Use through the Measurement of Sewage Drug Biomarkers. Environ Sci Technol. 2013 Feb 5;47(3):1452–60. doi:10.1021/es302722f

65. Ort C, Lawrence MG, Rieckermann J, Joss A. Sampling for Pharmaceuticals and Personal Care Products (PPCPs) and Illicit Drugs in Wastewater Systems: Are Your Conclusions Valid? A Critical Review. Environ Sci Technol. 2010 Aug 15;44(16):6024–35. doi:10.1021/es100779n

66. Gunduz I, Kondylis A, Jaccard G, Renaud JM, Hofer R, Ruffieux L, et al. Tobacco-specific N-nitrosamines NNN and NNK levels in cigarette brands between 2000 and 2014. Regulatory Toxicology and Pharmacology. 2016 Apr 1;76:113–20. doi:10.1016/j.yrtph.2016.01.012

67. Hecht SS, Hatsukami DK. Smokeless Tobacco and Cigarette Smoking: Chemical Mechanisms and Cancer Prevention. Nat Rev Cancer. 2022 Mar;22(3):143–55. doi:10.1038/s41568-021-00423-4 PubMed PMID: 34980891; PubMed Central PMCID: PMC9308447.

68 Products C for T. Results from the Annual National Youth Tobacco Survey (NYTS). FDA [Internet]. 2025 Jul 22 [cited 2026 Jun 4]. Available from: https://www.fda.gov/tobacco-products/youth-and-tobacco/results-annual-national-youth-tobacco-survey-nyts

69. Kimmel SE, Berlin JA, Miles C, Jaskowiak J, Carson JL, Strom BL. Risk of acute first myocardial infarction and use of nicotine patches in a general population. Journal of the American College of Cardiology. 2001 Apr 1;37(5):1297–302. doi:10.1016/S0735-1097(01)01124-X

70. Sia CH, Ko J, Zheng H, Ho AFW, Foo D, Foo LL, et al. Association between smoking status and outcomes in myocardial infarction patients undergoing percutaneous coronary intervention. Sci Rep. 2021 Mar 19;11:6466. doi:10.1038/s41598-021-86003-w PubMed PMID: 33742073; PubMed Central PMCID: PMC7979717.

71 Guidance to Reduce the Cardiovascular Burden of Ambient Air Pollutants: A Policy Statement From the American Heart Association | Circulation [Internet]. [cited 2026 Jul 30]. Available from: https://www.ahajournals.org/doi/10.1161/CIR.0000000000000930

72. Shao XT, Zhao YT, Jiang B, Li YY, Lin JG, Wang DG. Evaluation of Three Chronic Diseases by Selected Biomarkers in Wastewater. ACS EST Water. 2023 Apr 14;3(4):943–53. doi:10.1021/acsestwater.2c00452

73. Wurtzler E, Barnell EK, Morrison C, Grass C, DuPré N, Biddle DJ, et al. Using wastewater for population-level colorectal cancer surveillance: a future research agenda. J Epidemiol Community Health. 2026 Mar 17. doi:10.1136/jech-2025-224253 PubMed PMID: 41844287.

74. Xiao A, Wu F, Bushman M, Zhang J, Imakaev M, Chai PR, et al. Metrics to relate COVID-19 wastewater data to clinical testing dynamics. Water Res. 2022 Apr 1;212:118070. doi:10.1016/j.watres.2022.118070 PubMed PMID: 35101695; PubMed Central PMCID: PMC8758950.

75. Zhang X, Lei Z, Wang Q, Wang R, Luo J, Jin E, et al. Comparative Performance of Wastewater, Clinical, and Digital Surveillance Indicators for COVID-19 Monitoring in Routine Practice: Retrospective Observational Study. J Med Internet Res. 2025 Nov 6;27:e70232. doi:10.2196/70232 PubMed PMID: 41200904; PubMed Central PMCID: PMC12592968.

76. Amin V, Bowes DA, Halden RU. Systematic scoping review evaluating the potential of wastewater-based epidemiology for monitoring cardiovascular disease and cancer. Sci Total Environ. 2023 Feb 1;858(Pt 3):160103. doi:10.1016/j.scitotenv.2022.160103 PubMed PMID: 36370774; PubMed Central PMCID: PMC9643312.

77. Kondo T, Nakano Y, Adachi S, Murohara T. Effects of Tobacco Smoking on Cardiovascular Disease. Circulation Journal. 2019;83(10):1980–5. doi:10.1253/circj.CJ-19-0323

78. Roy A, Rawal I, Jabbour S, Prabhakaran D. Tobacco and Cardiovascular Disease: A Summary of Evidence. In: Prabhakaran D, Anand S, Gaziano TA, Mbanya JC, Wu Y, Nugent R, editors. Cardiovascular, Respiratory, and Related Disorders [Internet]. 3rd ed. Washington (DC): The International Bank for Reconstruction and Development / The World Bank; 2017 [cited 2026 May 6]. Available from: http://www.ncbi.nlm.nih.gov/books/NBK525170/ PubMed PMID: 30212086.

79. Cigarette Smoking and Competing Risks for Fatal and Nonfatal Cardiovascular Disease Subtypes Across the Life Course | Journal of the American Heart Association [Internet]. [cited 2026 May 6]. Available from: https://www.ahajournals.org/doi/10.1161/JAHA.121.021751

80. Wu X, Zhang H, Qi W, Zhang Y, Li J, Li Z, et al. Nicotine promotes atherosclerosis via ROS-NLRP3-mediated endothelial cell pyroptosis. Cell Death Dis. 2018 Feb 7;9(2):171. doi:10.1038/s41419-017-0257-3

81. Messner B, Bernhard D. Smoking and Cardiovascular Disease. Arteriosclerosis, Thrombosis, and Vascular Biology. 2014 Mar;34(3):509–15. doi:10.1161/ATVBAHA.113.300156

82. Kotlyarov S. The Role of Smoking in the Mechanisms of Development of Chronic Obstructive Pulmonary Disease and Atherosclerosis. Int J Mol Sci. 2023 May 13;24(10):8725. doi:10.3390/ijms24108725 PubMed PMID: 37240069; PubMed Central PMCID: PMC10217854.

83. Buccafusco JJ, Terry AV. The potential role of cotinine in the cognitive and neuroprotective actions of nicotine. Life Sciences. 2003 May 16;72(26):2931–42. doi:10.1016/S0024-3205(03)00226-1

84. Tanner JA, Novalen M, Jatlow P, Huestis MA, Murphy SE, Kaprio J, et al. Nicotine Metabolite Ratio (3-hydroxycotinine/cotinine) in Plasma and Urine by Different Analytical Methods and Laboratories: Implications for Clinical Implementation. Cancer Epidemiol Biomarkers Prev. 2015 Aug;24(8):1239–46. doi:10.1158/1055-9965.EPI-14-1381 PubMed PMID: 26014804; PubMed Central PMCID: PMC4526326.

85. von Weymarn LB, Lu X, Thomson NM, LeMarchand L, Park SL, Murphy SE. Quantitation of ten urinary nicotine metabolites, including 4-hydroxy-4-(3-pyridyl)butanoic acid, a product of nicotine 2’-oxidation, and CYP2A6 activity in Japanese Americans, Native Hawaiians and Whites. Chem Res Toxicol. 2023 Feb 20;36(2):313–21. doi:10.1021/acs.chemrestox.2c00413 PubMed PMID: 36735658; PubMed Central PMCID: PMC10042446.

86. Lin W, Huang Z, Gao S, Luo Z, An W, Li P, et al. Evaluating the stability of prescription drugs in municipal wastewater and sewers based on wastewater-based epidemiology. Science of The Total Environment. 2021 Feb 1;754:142414. doi:10.1016/j.scitotenv.2020.142414

87. Baker DR, Kasprzyk-Hordern B. Critical evaluation of methodology commonly used in sample collection, storage and preparation for the analysis of pharmaceuticals and illicit drugs in surface water and wastewater by solid phase extraction and liquid chromatography–mass spectrometry. Journal of Chromatography A. 2011 Nov 4;1218(44):8036–59. doi:10.1016/j.chroma.2011.09.012

88. Burgard DA, Williams J, Westerman D, Rushing R, Carpenter R, LaRock A, et al. Using Sewers to Monitor the Effects of Legalized Retail Sales on Cannabis Consumption in Washington State, USA. Addiction. 2019 Sep;114(9):1582–90. doi:10.1111/add.14641 PubMed PMID: 31211480; PubMed Central PMCID: PMC6814135.

89. Humphries MA, Bruno R, Lai FY, Thai PK, Holland BR, O’Brien JW, et al. Evaluation of Monitoring Schemes for Wastewater-Based Epidemiology to Identify Drug Use Trends Using Cocaine, Methamphetamine, MDMA and Methadone. Environ Sci Technol. 2016 May 3;50(9):4760–8. doi:10.1021/acs.est.5b06126

